# Spike-specific T cells are enriched in breastmilk following SARS-CoV-2 mRNA vaccination

**DOI:** 10.1101/2021.12.03.21267036

**Authors:** Blair Armistead, Yonghou Jiang, Marc Carlson, Emily S Ford, Saumya Jani, John Houck, Xia Wu, Lichen Jing, Tiffany Pecor, Alisa Kachikis, Winnie Yeung, Tina Nguyen, Nana Minkah, Sasha E Larsen, Rhea N Coler, David M Koelle, Whitney E Harrington

## Abstract

Human breastmilk is rich in T cells; however, their specificity and function are largely unknown. We compared the phenotype, diversity, and antigen specificity of T cells in the breastmilk and peripheral blood of lactating individuals who received SARS-CoV-2 mRNA vaccination. Relative to blood, breastmilk contained higher frequencies of T effector and central memory populations that expressed mucosal-homing markers. T cell receptor (TCR) sequence overlap was limited between blood and breastmilk. Overabundant breastmilk clones were observed in all individuals, were diverse, and contained CDR3 sequences with known epitope specificity including to SARS-CoV-2 Spike. Spike-specific TCRs were more frequent in breastmilk compared to blood and expanded in breastmilk following a third mRNA vaccine dose. Our observations indicate that the lactating breast contains a distinct T cell population that can be modulated by maternal vaccination with potential implications for infant passive protection.

**One-Sentence Summary:** The breastmilk T cell repertoire is distinct and enriched for SARS-CoV-2 Spike-specificity after maternal mRNA vaccination.

## Introduction

The breastfed human infant consumes up to 750,000 maternal leukocytes per day, 5 -10% of which are T cells, the function of which is poorly understood (*1, 2*). Breastmilk lymphocytes are most abundant at delivery and decline over the first month post -partum to a steady state that persists for up to two years. (*1-4*). However, the infant’s exposure to breastmilk cells likely remains substantial throughout breastfeeding due to an increase in volume of breastmilk ingested as the infant grows (*5*). Breastmilk T cells are phenotypically distinct from peripheral blood T cells, with higher expression of mucosal and effector memory markers (*6, 7*). Cytometaglovirus (CMV), Epstein-Barr virus (EBV), influenza, and HIV-specific T cell responses have been detected in breastmilk cells (BMC) at higher frequencies than in peripheral blood mononuclear cells (PBMC) (*7-11*), and breastmilk T cells expand in the setting of maternal or infant infection (*2, 12, 13*).

The infant stomach pH (*14, 15*) and intestinal permeability (*14, 16, 17*) are also highest in the first few weeks of life, and evidence from animal models demonstrates that breastmilk T cells can survive the offspring gastrointestinal tract and traffic into the mesenteric lymph nodes, liver, spleen, and lung as a form of maternal microchimerism (*18-20*). In mice, breastmilk-derived helminth-specific T cells were protective in the offspring upon challenge with the same helminth (*18*), and in lambs, breastmilk-derived tetanus-specific T cells enhanced the response to tetanus vaccination in the offspring (*21*). Human breastmilk maternal microchimerism has not been conclusively demonstrated, although we recently found in a cohort of infants th at maternal microchimerism increased up to three months of age and was positively associated with breastfeeding (*22*). These data emphasize the potential for breastmilk-acquired maternal T cells to become resident in the infant and provide an underrecognized form of passive protection.

The full repertoire of breastmilk T cells has not been described, however, and it is unclear whether maternal vaccine-specific T cells are present in human breastmilk. In the setting of the ongoing pandemic, pregnant people are now widely receiving SARS-CoV-2 (SARS2) vaccines including the Spike protein mRNA-based vaccines mRNA1273 (Moderna) (*23*) and BNT162b2 (Pfizer-BioNTech) (*24*). SARS2 mRNA vaccines generate a robust T cell response in peripheral blood (*23-26*) and induce the expansion of tissue resident T cell populations in the respiratory mucosa (*27*), yet their impact on mucosal T cell responses in breastmilk has been little studied (*28*). To understand the breadth of maternal T cells consumed by the infant, we characterized the phenotype and diversity of paired breastmilk and peripheral T cells. We further investigated the hypothesis that Spike-specific T cells are present in the breastmilk of SARS2 mRNA-vaccinated individuals and expand upon antigen re-encounter.

## Results

### Breastmilk is enriched for mucosal memory T cells

We collected paired blood and breastmilk from lactating people who had received two doses of BNT162b2 or mRNA1273 and had no history of SARS2 infection (Fig. S1; Table 1). We characterized cell phenotype by flow cytometry in paired samples with at least 1,000,000 total BMC recovered and at least 100 cells in both the CD4 + and CD8+ T cell populations (n=17). Breastmilk contained a low but detectible frequency of T cells, and the frequency of CD4+ T cells was similar in BMC and PBMC, whereas the frequency of CD8+ T cells was somewhat lower in BMC than PBMC (24% vs. 30%, p=0.03; Fig. S2). Next, we identified naïve and memory T cell subsets in BMC and PBMC through surface expression of CD45RO (marker of antigen experience) and CCR7 (chemokine receptor that facilitates homing to secondary lymphoid organs). We found a significant enrichment of CD45RO+/CCR7-effector memory (T_EM_) (84% vs. 41%, p<0.001) and CD45RO+/CCR7+ central memory (T_CM_) CD4+ T cell populations (11% vs. 6%, p=0.006) in BMC vs. PBMC (Fig. S3A, B). Within the CD8+ population, there was also a higher frequency of T_EM_ (73% vs. 38%, p<0.001) but not T_CM_ (1% vs. 1%, p=0.9) in BMC relative to PBMC (Fig. S3A, B). The ratio of T_EM_ to T_CM_ frequencies was consistent in BMC and PBMC (CD4+: 10 vs. 9, p=0.4; CD8+: 85 vs. 63, p=0.2). These data emphasize that breastmilk is highly enriched for memory T cell populations relative to peripheral blood.

**Table 1.** Cohort characteristics.

|  | **Total (n=20)** |
| --- | --- |
| **Maternal age, years, median (range)** | 37 (29-39) |
| **Maternal vaccine type, n (%)** |  |
| Pfizer-BioNTech | 17 (85%) |
| Moderna | 3 (15%) |
| **Time since delivery, weeks, median (range)** | 26 (2-56) |
| **Time since second SARS2 mRNA dose, weeks, median (range)** | 11 (0.7-24) |
| **Parity, median (range)** | 1.5 (1-3) |
| **Infant sex, n (%)** |  |
| Female | 7 (33%) |
| Male | 14 (67%) |

We next investigated the expression of mucosal-homing markers CCR9 and CD103 on T cells within BMC and PBMC. The CD4+ population within BMC versus PBMC had a higher frequency of CCR9+ (38% vs. 4%, p<0.001) and CD103+ (7% vs. 0.4%, p<0.001) cells, and a higher frequency of double positive CCR9+/CD103+ cells (5% vs. 0.1%, p<0.001) (Fig. S4A, B). Similarly, the CD8+ population within BMC versus PBMC had a higher frequency of CCR9+ (12% vs. 3%, p=0.005) and CD103+ (32% vs. 3%, p<0.001) cells, as well as a higher frequency of CCR9+/CD103+ cells (3% vs. 0.4%, p<0.001) (Fig. S4A, B). These data emphasize that the T cells in breastmilk express high levels of mucosal-homing markers.

### Restricted T cell receptor repertoire overlap between peripheral blood and breastmilk

Because of the high frequency of memory populations, we next investigated the composition and diversity of the T cell repertoire (TCR) in BMC. BMC were enriched for hematopoietic lineage cells (CD45+) using flow cytometry cell sorting. Sorted BMC for which >1,000 DNA genomic equivalents were recovered (n=16) underwent bulk T cell receptor beta chain (TCRβ) sequencing. Among BMC samples for which >1,000 TCR templates were recovered, we compared the degree of TCR overlap with matched peripheral blood (n=11) and surprisingly observed relatively low overlap in all pairs, as measured by the Morisita and power geometric indices (*29*) (Fig. 1, File S1). In contrast, we observed a high degree of overlap in the PBMC TCR repertoire from one individual at two time points (9 and 17 days post-2nd vaccine dose) utilized as a positive control (Fig. 1, File S1). Neither the Morisita index nor the power geometric index between paired BMC and PBMC was related to the number of productive templates in the BMC (R^2^=0.4, p=0.3; R^2^=0.3, p=0.4 respectively), suggesting that the low overlap was independent of sampling depth of the BMC. Within each participant, we compared the frequency of clonotypes across the two compartments using the immunoSEQ® Differential Abundance tool (*30*). In all individuals, there were select clonotypes that were statistically significantly overabundant in BMC relative PBMC (Fig. 1; File S1). In contrast, the Simpson Clonality and maximum clone frequency—two metrics of absolute clonality—did not differ in BMC and PBMC. These data indicate that the TCR repertoire of BMC is highly diverse at the sequence level with select overabundant clonotypes compared to PBMC.

**Fig. 1.**
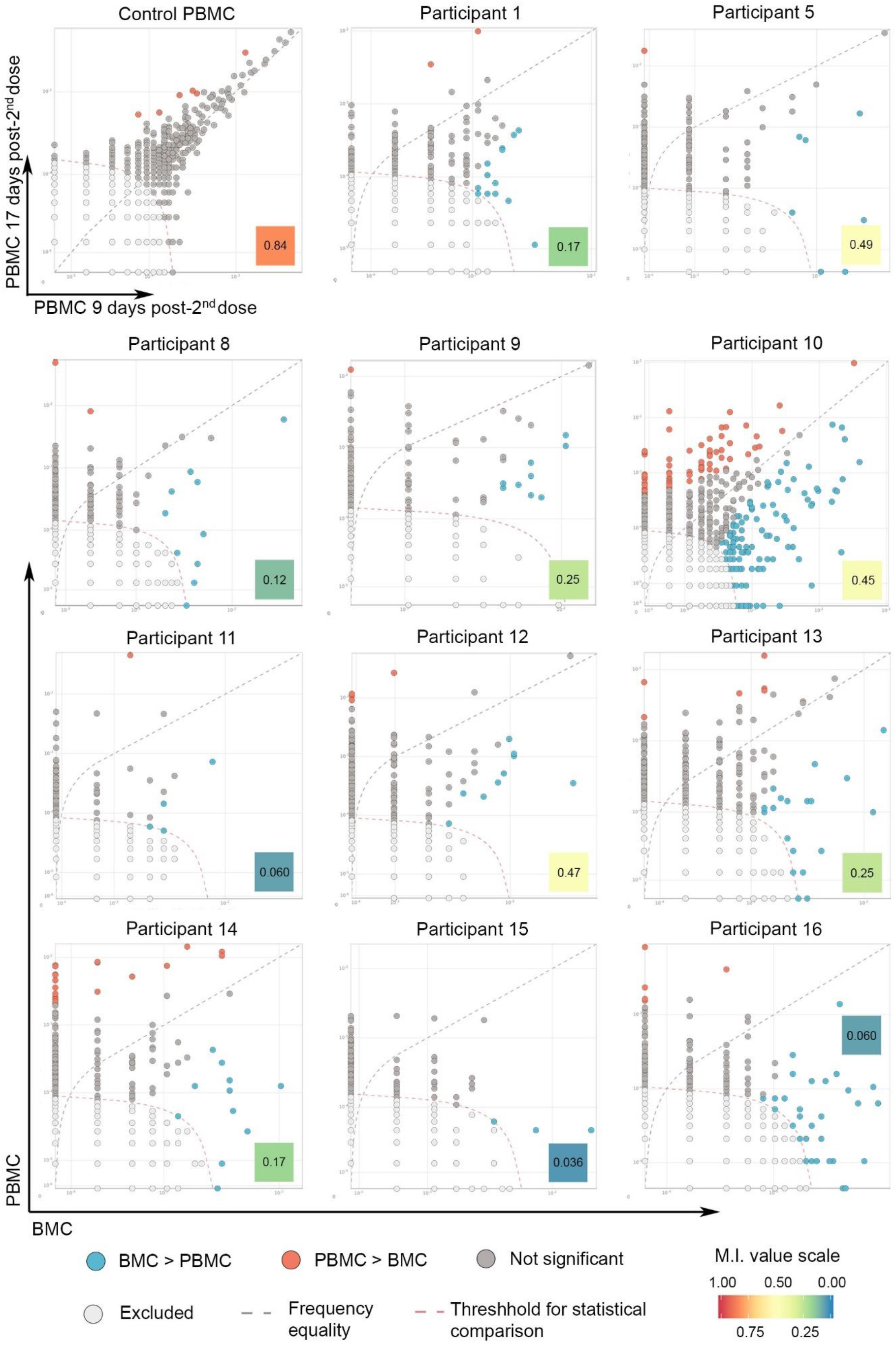
The T cell receptor (TCR) repertoires in breastmilk and peripheral blood are distinct. Bulk TCRβ sequencing from BMC and PBMC individuals who had paired samples available and at least 1,000 TCRβ templates in the BMC sample (n=11). The frequencies of TCRβ clonotypes in the two compartments were compared using the immunoSEQ® Differential Abundance Tool. TCRβ repertoire overlap was analyzed using the Morisita index (M.I., value inset). As a control, TCRβ clonotypes from an individual’s PBMCs obtained 9 days and 17 days after 2^nd^ mRNA vaccine dose were compared (upper left plot).

### Characterization of breastmilk overabundant T cell clonotypes

We next explored the diversity of the overabundant T cell clonotypes in breastmilk. We analyzed each participant’s BMC T cell repertoire using a TCR distance metric, tcrdist3 (*31, 32*), which clusters TCRβs based on structural and functional similarities of amino acids within complementarity-determining regions (CDR). In all participants, overabundant clones were broadly distributed across the BMC TCRβ repertoire (Fig. 2A, Fig. S5). We additionally evaluated how overabundant clones clustered across all individuals. Most clones were unique to a single individual (i.e., they were “private”), and the clonotypes did not segregate by individual (Fig. 2B), reflecting the diversity of each participant’s overabundant clonotypes.

**Fig. 2.**
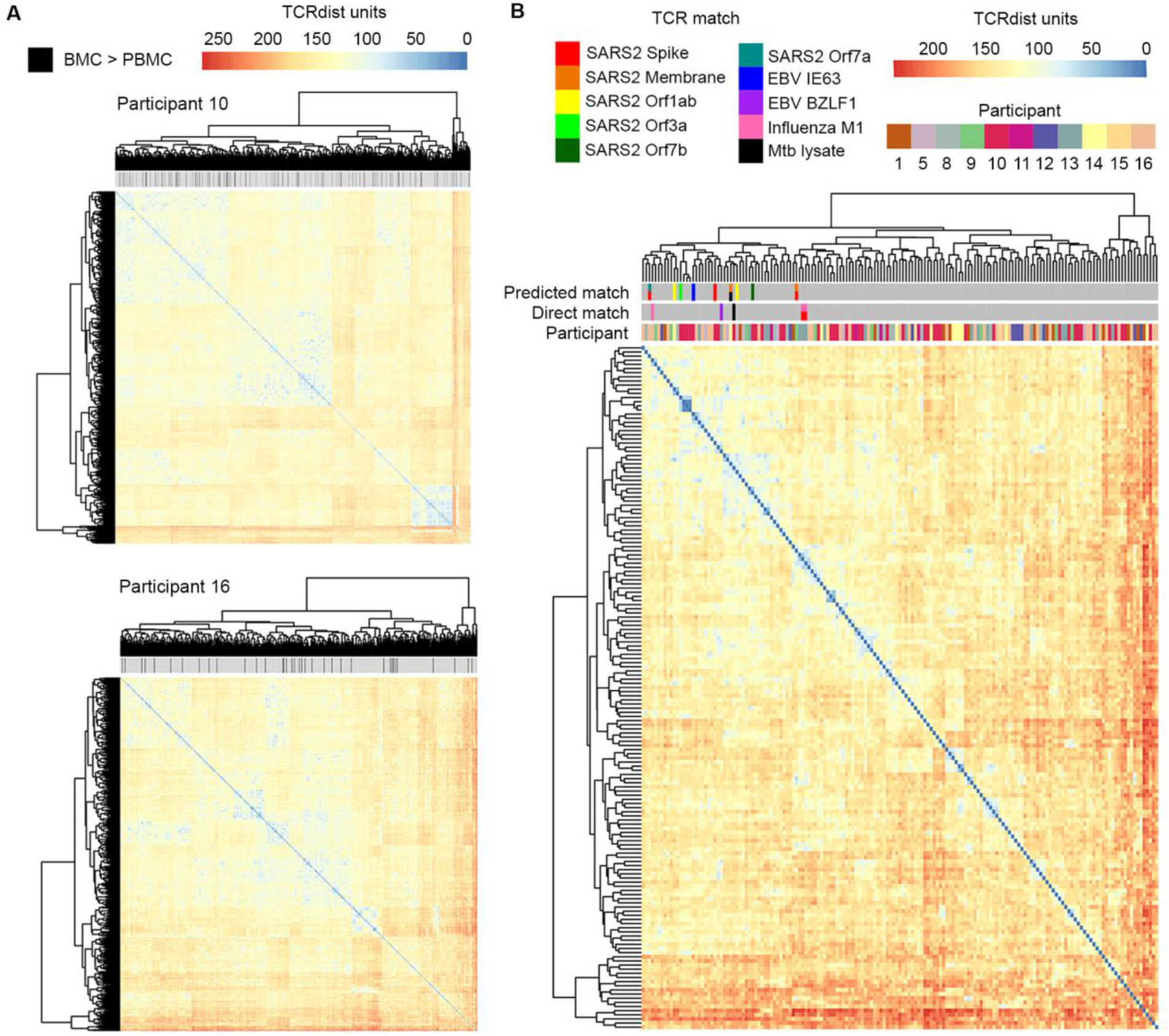
Overabundant TCR clones in breastmilk are diverse. (**A**) All TCRβ CDR3 amino acid sequences obtained from the BMC of each participant for which paired PBMC was available (n=11) were compared using tcrdist3; representative plots from two individuals are displayed. Black ticks denote TCRβ sequences significantly overabundant in BMC relative to PBMC. (**B**) Overabundant BMC TCRβ CDR3 amino acid sequences from 11 participants were compared to one another using tcrdist3. Overabundant TCRβ clones were compared by CDR3 amino acid sequence and V gene usage against available public databases of known TCR epitope specificity. Clones matching pathogen-specific epitopes are marked with colored ticks. For Participant 10, only CDR3 amino acid sequences enriched by a factor ≥50 relative to PBMC or with epitope specificity were included to reduce data skewing from this participant.

To determine potential antigen specificity, we compared CDR3 amino acid sequences of the overabundant breastmilk clones from all individuals to TCRβ sequence databases populated by validated epitope-specific TCRs. We identified five direct matches from two participants with CDR3 amino acid sequences and identical V gene usage, two of which were reported to bind SARS2 Spike epitopes (Fig. 2B, Table S1). Notably, the two Spike-specific clones had also previously been reported to bind influenza M1 (*33, 34*). Other direct matches included epitopes derived from influenza, *Mycobacterium tuberculosis* lysate, and EBV (Fig. 2B, Table S1).

Additional clones with identical CDR3 sequences but non-identical V gene usage were reported to bind epitopes from SARS2, EBV, and *M. tuberculosis* (Fig. 2B, Table S1). CDR3 sequences with previously published specificity clustered tightly together, irrespective of individual (Fig 2B). Finally, we used the IEDB TCRMatch Tool (*35*) to predict TCR-epitope specificity based on sequence similarity to published TCR sequences, which identified TCR clones with potential specificity to a variety of viral epitopes (Table S2). These data indicate that overabundant breastmilk T cell clones are diverse and respond to a range of pathogen-specific epitopes.

### SARS2 Spike-restricted TCRβs are present in breastmilk T cells

Because all participants had received SARS2 Spike mRNA vaccination, we next investigated the presence of Spike-specific clones in BMC T cells more broadly. We utilized the immunoSEQ® COVID Search Tool (*36*) to identify candidate Spike-specific TCRβ in all sequenced BMC and PBMC samples (n=30). All PBMC (n=14) contained candidate Spike-specific TCRβ, though their predicted epitope specificity was distributed across the entire Spike protein with low frequency, suggesting that some of the TCRβ may represent clones in the naïve repertoire rather than expanded vaccine-specific populations (Fig. 3A). Thirteen of the 16 BMC samples contained TCRβ predicted to be Spike-specific. In the pairs where both BMC and PBMC were sampled and at least one clone in each compartment was predicted to be SARS2 -specific (n=13 pairs), Spike-specific TCRβs were nearly 2-fold enriched in BMC versus PBMC (Fig. 3B). Notably, the frequency of Spike-specific TCRβs were not strongly correlated with Spike-specific IgG or IgA in breastmilk or blood (Fig. S6).

**Fig. 3.**
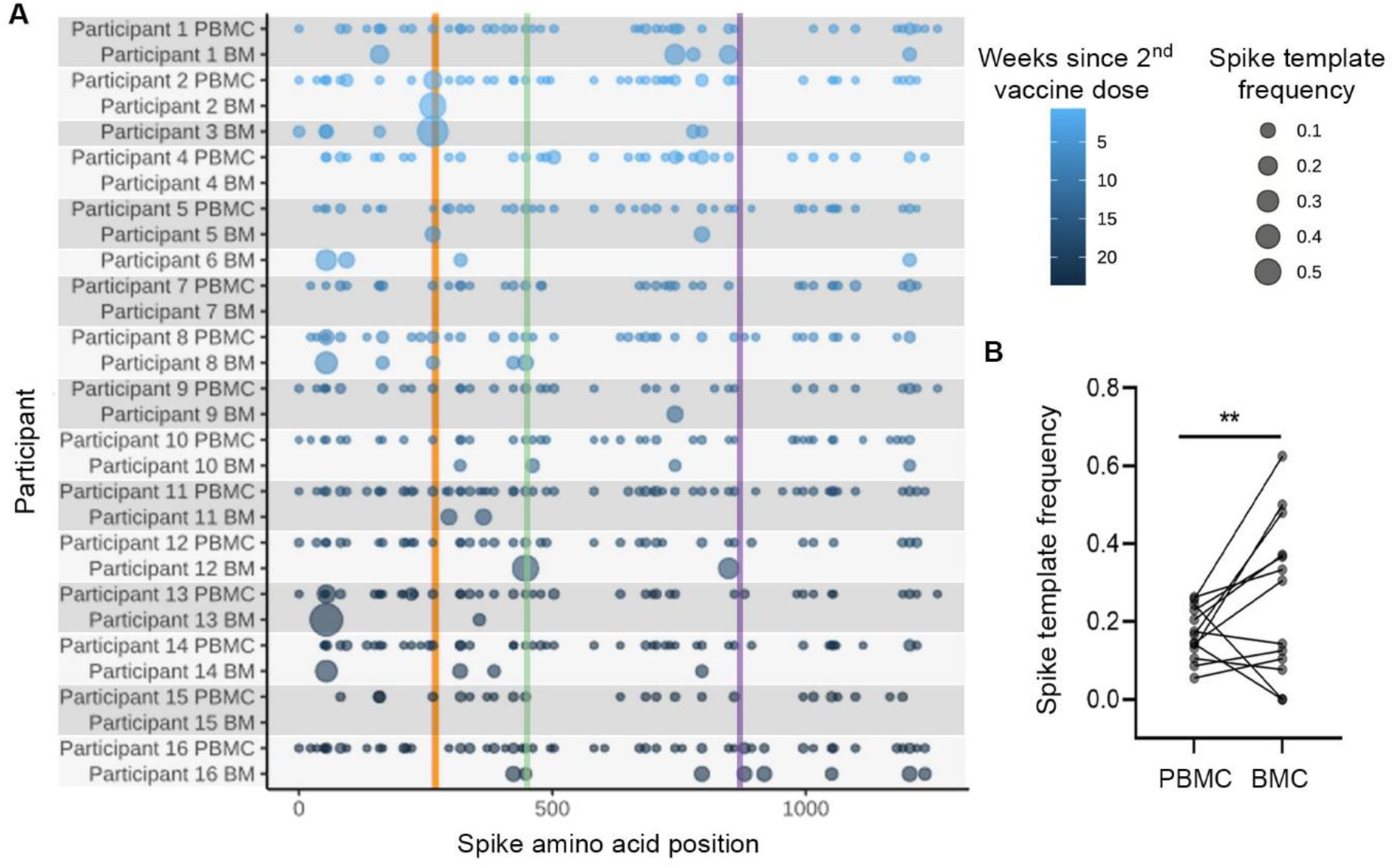
Candidate SARS2 Spike-specific T cells are enriched in the breastmilk relative to the peripheral blood of vaccinated individuals. The frequency of candidate Spike-specific clones in all sequenced PBMC and BMC (n=14 paired, n=2 BMC only) is expressed relative to all clones predicted to bind to SARS2 antigens. Spike template frequency was compared using a negative binomial model, **p<0.01. (**A**) TCRβ sequences predicted to bind to SARS2 were identified in BMC and PBMC using the ImmunoSEQ® T-MAP COVID Search Tool and are mapped by their epitope binding location on the Spike protein. Gold line indicates the amino acid position of the Spike peptide pool that includes the YLQPRTFLL epitope, green line indicates the position of the NYNYLYRLF epitope, and purple line indicates the positio n of the LTDEMIAQY epitope. (**B**) Spike-specific TCRβ templates are enriched in BMC relative to PBMC, incident rate ratio (IRR)=166, p=0.004.

**Fig. 4.**
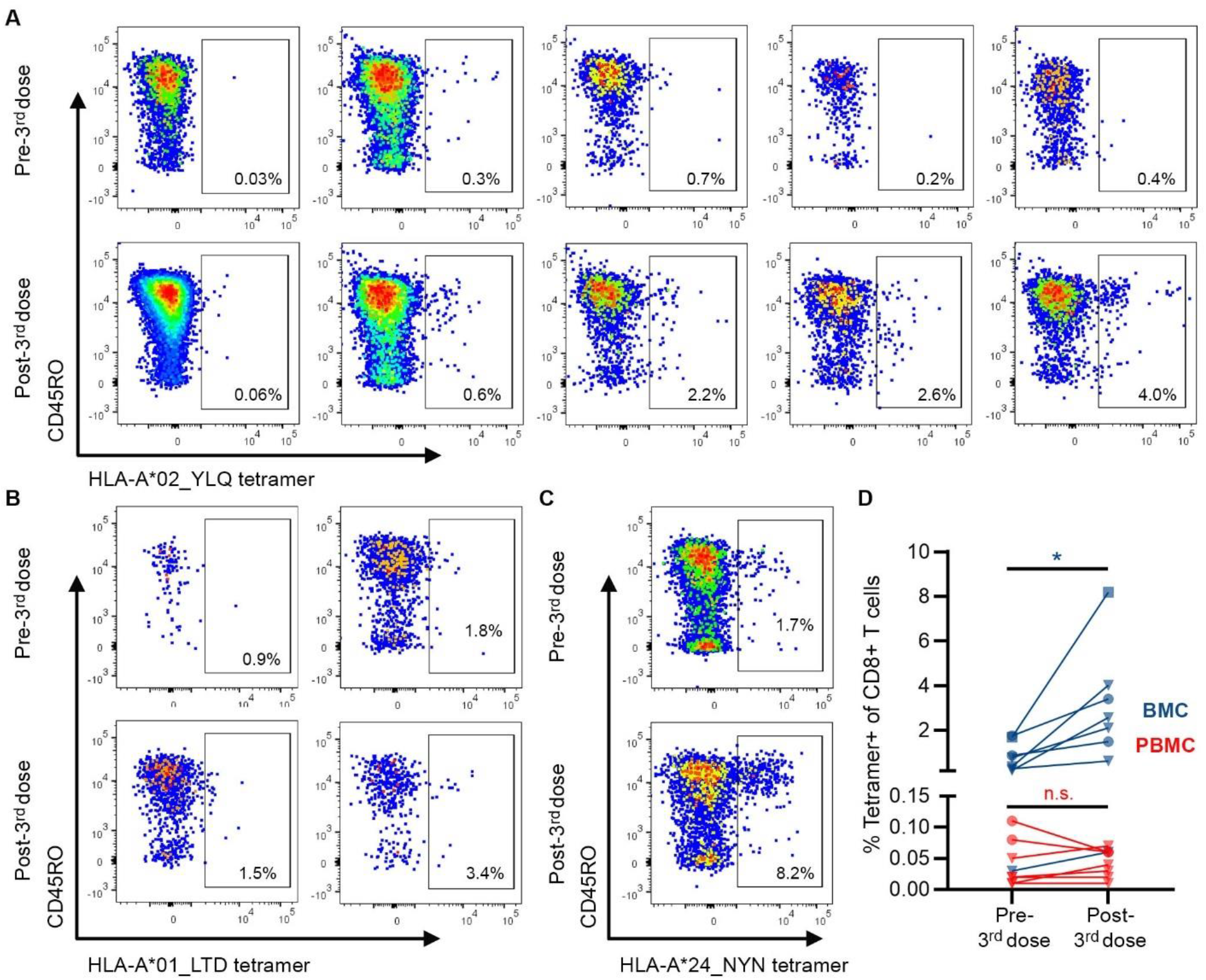
SARS2 Spike-specific T cells expand in breastmilk following 3^rd^ mRNA vaccinedose. BMC and PBMC from before and after receipt of the 3^rd^ dose of SARS2 mRNA vaccine were stained with SARS2 Spike epitope-loaded class I tetramers and analyzed by flow cytometry to quantify Spike-specific CD8+ T cells. Comparisons made with paired t tests, *p<0.05, n.s. (non-significant). Scatter plots of HLA-A*02_YLQ-positive (n=5) (**A**), HLA-A*01_LTD-positive (n=2) (**B**), and HLA-A*24_NYN-positive (n=1) (**C**) CD8+ T cells in breastmilk obtained before (top) and after (bottom) 3^rd^ mRNA vaccine dose are shown from HLA concordant individuals. Frequencies of tetramer+ cells of CD8+ T cells inset. (**D**) Frequencies of tetramer+ cells of CD8+ T cells in BMC (n=8, blue) and PBMC (n=7, red). Triangle=HLA-A*02_YLQ, circle=HLA-A*01_LTD, square=HLA-A*24_NYN. BMC: 0.8% to 2.8%, p=0.03; PBMC: 0.04% to 0.04%, p=0.9.

To further validate the presence of Spike-specific T cells in breastmilk, we cross-referenced each participant’s BMC and PBMC TCRβ CDR3 amino acid sequences and V gene usage against publicly available TCRβ datasets from Spike-epitope-loaded tetramer or multimer experiments (*37-41*). We identified high quality CDR3 sequence matches in half of all breastmilk samples, with five samples also containing hits with identical V gene usage (Table S3). All PBMC contained clones with identical CDR3 sequence and V gene usage to those published previously (Table S4). Consistent with prior studies of PBMC (*37-39, 42*), sequences specific to the Spike peptide YLQPRTFLL were prominent in the breastmilk and blood of individuals known or presumed to be HLA-A*02:01 positive. In addition, clones specific to the A*01:01 -restricted Spike peptide LTDEMIAQY and the B*15:01-restricted Spike peptide NQKLIANQF were present in the breastmilk and blood of HLA concordant participants (Table S3, Table S4), suggesting shared epitope specificity following vaccination. Finally, we utilized the list of Spike -specific TCRβs in conjunction with the tcrdist3 algorithm (*31, 32*) to identify novel potential Spike-specific T cell clones in the BMC (Table S5). These observations demonstrate the presence of Spike-specific T cells in breastmilk following mRNA vaccination.

### Spike-specific T cells in breastmilk expand after SARS2 mRNA vaccine

To understand whether Spike-specific clones in breastmilk were responsive to antigen re-exposure, we took advantage of a natural restimulation experiment in which participants (n=8) donated breastmilk pre- and approximately 1-week post-3rd vaccine dose for in-depth phenotyping and staining with HLA class I tetramers loaded with immunodominant Spike epitopes, namely HLA-A*02_YLQ, HLA-A*01_LDT, and HLA-A*24_NYN (Table 2, Fig. 6A, B, C) (*37-39, 42*). The proportion of Spike tetramer-positive CD8+ T cells of all CD8+ T cells significantly increased in breastmilk between the pre- and post-3^rd^ dose samples (0.8% to 2.8%, p=0.03), whereas tetramer-positive CD8+ T cells in PBMC showed minimal response (0.04% to 0.04%, p=0.9) (Fig. 6D). The expression of activation markers on tetramer+ CD8+ T cells was consistently high but did not vary between pre- and post-3^rd^ dose samples including CD69 (51% vs. 50%, p=0.9), CD137 (61% vs. 42%, p=0.3), and CCR5 (89% vs. 84%, p=0.4). The proportion of tetramer+ CD8+ T cells expressing CCR9 and CD103 was similar to our earlier bulk analysis and did not vary between pre- and post-3^rd^ dose (CCR9: 50% vs. 31%, p=0.3; CD103: 18% vs. 7%, p=0.4; CCR9/CD103: 1.4% vs. 3.4%, p=0.3). A proportion of tetramer+ cells also expressed the lymphocyte integrin α4β7, which regulates T cell migration to the intestine (21% vs. 16%, p=0.7). These data demonstrate that SARS2 Spike-specific cells in breastmilk respond *in vivo* upon antigen restimulation.

**Table 2.** Spike epitope-loaded tetramers used in this study.

| **Tetramer**<br>**annotation in text** | **SARS2 Spike**<br>**epitope**<br>**sequence** | **Sequence**<br>**position within**<br>**Spike protein** | **HLA class I**<br>**restriction** | **Conjugated**<br>**fluorophore** |
| --- | --- | --- | --- | --- |
| HLA-A*02_YLQ | YLQPRTFLL | 269-277 | HLA-A*02:01 | APC |
| HLA-A*24_NYN | NYNYLYRLF | 448-456 | HLA-A*24:02 | APC |
| HLA-A*01_LTD | LTDEMIAQY | 865-873 | HLA-A*01:01 | APC |

## Discussion

We present a comprehensive comparison of the T cells present in breastmilk relative to peripheral blood, considering phenotype, diversity, and antigen specificity. We find that T cells in breastmilk are nearly uniformly memory populations and have high expression of mucosal -homing markers. Their TCRβ repertoire is diverse, yet distinct from paired PBMC, and with select overabundant clones. Further, we identify SARS2 Spike-specific clones in the breastmilk of mRNA vaccinated individuals, emphasizing that vaccine-specific T cells are present at mucosal sites such as the breast, with important implications for both maternal and infant health.

In bulk analysis, breastmilk T cells were enriched for effector memory (i.e., T_EM_) populations, indicating that breastmilk T cells may be poised to rapidly respond following antigen re-encounter (*43*). Further, breastmilk T cells displayed high levels of mucosal-homing markers, consistent with earlier reports in individuals living with or without HIV (*7*). These data suggest that breastmilk T cells may be derived from a tissue-resident population in the breast (*44*), rather than due to vessel microtrauma and contamination by peripheral blood. The high expression of both CCR9 and CD103 by breastmilk T cells also supports the notion of an entero-mammary axis (*45*). Future studies should investigate whether breastmilk T cells traffic to the infant respiratory and gastrointestinal tracts when consumed and enhance cellular immunity to pathogens, as has been shown in helminthic infection in mice (*18*), or amplify infant response to homologous vaccination, as has been shown with tetanus vaccination in lambs (*21*).

The TCRβ repertoire in BMC was diverse and had uniquely expanded clonotypes relative to paired PBMC. To date, studies have focused on T cell responses to specific pathogens (*7-12*), rather than capturing the full diversity of the compartment. The low degree of TCRβ repertoire overlap between the BMC and PBMC may reflect a difference in the distribution of naïve versus antigen-experienced T cells. However, the absolute clonality was similar and there was evidence of high frequency clonotypes, likely memory cells, that were differentially abundant, suggesting distinct T cell responses in the two compartments independent of the naïve population. The observation of overabundant clones in the BMC is consistent with prior reports of an enrichment of virus-specific responses in breastmilk relative to PBMC (*7-10*), although in our paired TCRβ analysis only one individual had CMV-specific TCRβ and none reported HIV infection, emphasizing that this breast-specific enrichment is not restricted to virus-specific clones. In addition, within each individual, overabundant clones were diverse.

To identify antigen specificity of BMC T cells, we used a combination of prior published TCRβ specificities and predictive algorithms, which most notably allowed for the high -confidence detection of SARS2 Spike-specific clones in most individuals. Consistent with previous observations of convergent epitope specificity across HLA-concordant people following SARS2 mRNA vaccination (*37*), we found identical Spike-specific CDR3 sequences in BMC of several individuals (Table S3). This observation, along with the approval of 3^rd^ mRNA vaccine doses, provided a unique opportunity to observe the dynamics of Spike -specific CD8+ T cells in the breastmilk after antigen re-encounter using well-validated HLA Class I tetramers loaded with Spike epitopes (*37-39, 42*), including HLA-A*02_YLQ, HLA-A*01_LDT, and HLA-A*24_NYN (Table 2). In contrast to blood-derived CD8+ T cells, the frequency of tetramer-positive CD8+ T cells in breastmilk significantly increased after the 3^rd^ SARS2 mRNA vaccine dose, suggesting that T cells in the lactating breast may be particularly poised to respond to maternal mRNA vaccination. Tetramer+ CD8+ T cells in breastmilk had high expression of activation and mucosal homing markers, emphasizing their functional potential (*27*).

In addition to the potential benefit provided to the infant, the recognition of the breast as a site of mucosal immunity distinct from peripheral immunity has important implications for the study of vaccine responses. Prior work on the response to SARS2 mRNA vaccines has primarily focused on both CD4+ and CD8+ T cell responses present in peripheral blood (*23, 24*) with limited study of T cell responses at mucosal sites. One recent study found that SARS2 mRNA vaccination induces the expansion of resident CD8+ T cells in the upper respiratory tract (*27*), suggesting that that SARS2 mRNA vaccines promote robust T cell responses in the mucosa in addition to the periphery. In addition, a recent study reported an increase in Spike peptide-reactive T cells in breastmilk following mRNA vaccination (*28*), consistent with our observations. Notably, lower respiratory tract responses are difficult to access in human populations, whereas the collection of breastmilk in lactating individuals is non-invasive. While the association between respiratory tract and breast resident T cell responses will need further investigation, it is possible th at measuring immune responses in breastmilk will allow for the characterization of mucosal immunity more broadly following vaccination.

Our study had several limitations. BMC T cells were low frequency, and although we designed our experimental approach to maximize the information obtained from each sample, the sampling depth of BMC versus PBMC was different. However, we took advantage of several computational solutions to overcome this challenge, including repertoire analysis utilizing tools less susceptible to bias with differences in sampling depth (Morisita and power geometric indices) and down-sampling for clonality analysis. We were further limited to bulk TCRβ sequencing of combined CD4+ and CD8+ populations, as the number of each sub -population was too small to meet technical requirements to analyze separately. Future work should consider the use of single cell RNAseq to investigate the two populations separately. The number of T cells recovered from each breastmilk limited our ability to conduct functional analyses. However, we took advantage of individuals who received a 3^rd^ dose of SARS2 mRNA vaccine to demonstrate expansion of Spike-specific T cells with tetramer staining. We utilized previously frozen and thawed BMC and PBMC, which may have biased recovery of cell populations, particularly in the breastmilk. Future studies should consider detailed analysis of fresh BMC if possible. Finally, initiation and support for breastfeeding varies by demographic groups including socioeconomic status, and future work should address a more diverse population of lactating people.

We demonstrate that breastmilk T cells are highly diverse and enriched for mucosal memory populations, emphasizing that the lactating breast represents a key site of mucosal immunity that warrants additional study. Further, we identify SARS2 Spike-specific T cells in mRNA vaccinated individuals, a critical demonstration of vaccine-specific T cells in breastmilk. This observation may have important implications for both the study of vaccine-induced T cell responses in the vaccinated individual as well as for infant passive protection.

## Materials and Methods

### Cohort

The initial cohort was comprised of lactating individuals (n = 26, Fig. S1) who were recruited as part of the Center for Global Infectious Disease Research Biorespository, approved by the Seattle Children’s Institutional Review Board (IRB) (STUDY00002048). All participants self -reported as healthy, not pregnant, and weighing > 110 pounds. Demographic data was collected including age, history of SARS or other serious infections, SARS2 vaccination (brand of vaccine received, dates of doses), sex of infant(s), and parity. All participants reported receipt of two doses of a SARS2 Spike mRNA vaccine. No participants reported SARS2 infection, and all were negative for anti-nucleocapsid IgG by commercial testing (Abbott Architect SARS-CoV-2 IgG assay). Breastmilk was pumped by before the visit by the participants using the method of their choice and transferred fresh to the study team (not frozen). Blood was obtained from 23 individuals. For the pre- and post-3^rd^ mRNA vaccine dose studies, blood and breastmilk were collected before and approximately 1 week after 3^rd^ mRNA vaccine dose (n=7 PBMC + BMC, n=1 BMC only). Collection interval was based on T cell responses observed in peripheral blood (*23, 24*). Five participants from the original cohort returned to provide additional specimens, 1 new participant was enrolled, and samples from 2 additional individuals were added from the Maternal Immunizations in Low and High-Risk Pregnancies, approved by the University of Washington IRB (STUDY00008491). Participants from both studies provided written informed consent.

### Isolation of PBMCs from whole blood

Whole blood was collected in EDTA Vacutainer tubes (BD). Within 4 hours of blood collection, tubes were centrifuged at 400 x *g* for 10 minutes. The plasma fraction was removed, centrifuged at 800 x *g* for 15 minutes, aliquoted into cryovials, and stored at -80°C. The remaining blood was diluted in sterile phosphate buffered saline (PBS), layered onto Lymphocyte Separation Medium (Corning), and centrifuged at 800 x *g* for 20 minutes at room temperature with no brake. The resulting buffy coat layer was removed and washed two to three times in PBS. Cells were counted using a C-Chip hemocytometer (INCYTO), resuspended in freezing medium (50% fetal bovine serum [FBS], 40% RPMI with L-glutamine, 10% DMSO (Millipore Sigma)) at 5 to 10 million cells/mL, and aliquoted into cryovials. Cryovials were immediately placed into a 1°C cryogenic freezing container (Nalgene), which was stored at -80°C overnight. Cryovials were then moved to liquid nitrogen for long-term storage.

### Isolation of BMC

Milk was centrifuged at 400 x *g* for 15 minutes at 4°C, and the aqueous fraction was aliquoted into cryovials and stored at -80°C. The cell pellet was washed three times in 40 mL sterile PBS with 2% FBS. As above, cells were counted using a C-Chip hemocytometer (INCYTO), resuspended in freezing medium at 1 to 3 million cells/mL, aliquoted into cryovials, placed in a 1°C cryogenic freezing container at -80°C overnight, and then transferred to liquid nitrogen. Breastmilk samples with less than ∼10^6^ cells were not utilized for further analysis.

### Thawing PBMC and BMC

Prior to use in assays, PBMC or BMC were thawed in a 37°C water bath until a small ice crystal remained. Resuspensions were transferred into a tube containing pre-warmed thaw medium (RPMI with L-glutamine, 20% FBS) and centrifuged at 400 x *g* for 5 minutes. Cell pellets were resuspended in approximately 5 mL complete medium (RPMI with L-glutamine, 10% FBS, 100 U/mL penicillin, 100 μg/mL streptomycin) and then counted and assessed for viability before final resuspension in complete medium.

### Phenotyping and cell sorting by flow cytometry

PBMC and BMC were thawed as described and washed with FACS buffer (PBS, 2% BSA). Cells were stained with 100 μL of master mix comprised of the optimal dilutions of fluorophore-conjugated antibodies and viability dye (Table S6; panel 1) in Brilliant Stain Buffer (BD) for 30 minutes at room temperature, protected from light. Stained cells were washed with 4 mL FACS buffer and resuspended in 200 to 300 μL FACS buffer. Cells were run on a FACSMelody Cell Sorter (BD) or a FACSAria II Cell Sorter (BD), and CD45+ cells were collected. Single stained CompBeads (ThermoFisher) were used as compensation controls, and unstained or fluorescence-minus-one stained cells were used to set fluorescence gates. Data were analyzed on FlowJo version 10 (BD) and included analysis of memory markers and mucosal markers. All samples contained at least 100 cells in the CD4+ and CD8+ T cell subsets. CD45RO+/CCR7-T cells were designated as effector memory (T_EM_), CD45RO+/CCR7+ T cells were designated as central memory (T_CM_), and CD45RO-/CCR7+ T cells were designated as naïve-like (T_N-like_). Gating strategy is shown in Fig. S7. In addition, data were used to anticipate the frequency of T ce lls (CD3+) in the collected CD45+ population from BMC to optimize subsequent genomic DNA extraction.

### Tetramer generation

YLQPRTFLL (i.e. YLQ peptide), LTDEMIAQY (i.e. LTD peptide), and NYNYLYRLF (i.e. NYN peptide) were synthesized by GenScript Biotech (Piscataway, NJ). YLQ peptide (400 μM) was mixed 1:1 (v/v) with 200 μg/mL Flex-T HLA-A*02:01 ultraviolet exchange UVX monomer (BioLegend) and treated with UV irradiation (368 nm) for 30 minutes using a UV crosslinker (Fisher Scientific) to remove the UV-liable peptide. The mixture was incubated at 37°C for 30 minutes to form YLQ monomers, which were then tetramerized through the addition of 200 μg/mL streptavidin-APC (BD) and incubation on ice for 30 minutes. Excess streptavidin was blocked with PBS + 0.4 μM D-Biotin + 0.3% (w/v) NaN_3_ overnight at 4°C. HLA-A*01_LTD-APC and HLA-A*24_NYN-APC tetramers were generated by the National Institutes of Health Tetramer Core Facility (Emory University, Atlanta, GA).

### Tetramer staining and flow cytometry analysis

Cells were thawed and washed as described above and then stained with HLA-A*02_YLQ-APC, HLA-A*01_LTD-APC, or HLA-A*24_NYN-APC tetramer (1:100) for 30 minutes at 4°C, protected from light. Cells were washed with FACS buffer and then stained with antibody mix containing the optimal dilutions of all antibodies and viability dye in Brilliant Staining Buffer (Table S7; panel 2) for 30 minutes at room temperature, protected from light. As above, stained cells were washed in FACS buffer and run on a FACSAria II Cell Sorter (BD) with single stained CompBeads (ThermoFisher) as compensation controls. Data were analyzed on FlowJo version 10 (BD). HLA-A*02_YLQ tetramer performance was validated with YLQ-specific CD8+ T cells expanded from an HLA-A*02:01 SARS2 convalescent donor spiked into an HLA-A*02:01 negative donor with PBMC collected prior to 2019 (Fig. S8). HLA-A*01_LTD and HLA-A*24_NYN tetramer performance was validated using HLA-matched PBMC from donors predicted to have TCRs specific to these epitopes from TCRβ sequencing data and HLA-unmatched PBMC as negative controls. Tetramer positive cells from BMC and PBMC were gated as follows: lymphocyte cloud (FSC-A by SSC-A) ⟶ single cells (FSC-A by FSC-H) ⟶ live cells (live/dead Aqua by FSC-H) ⟶ CD45+/ CD3+ ⟶ CD8+ ⟶ tetramer+.

### HLA typing

Genomic DNA was extracted from participants’ PBMC (where available) with the QIAamp® DNA Blood Mini Kit per manufacturer instructions, and HLA class I genotyping was performed via direct sequencing (Sisco Genetics, Seattle, WA).

### TCRβ sequencing

Flow-sorted BMC (CD45+) expected to contain T cells underwent genomic DNA extraction using a protocol modified from that recommended by Qiagen to recover low-yield DNA. Briefly, sorted BMC were pelleted in their collection tubes at 400 x *g* for 10 minutes. Cell pellets were extracted directly in the collection tubes using 30 μL of Qiagen Protease (Qiagen) and incubated at 70°C for 20 minutes in a heat block with periodic vortexing. Approximately 1.5 μg of carrier RNA (Qiagen) and 300 μL of Buffer AL (Qiagen) was added. Samples were incubated for another 20 minutes at 70°C in a heat block with periodic vortexing and centrifugation. To recover DNA, 300 μL of 100% ethanol was added to the sample followed by vortexing. The sample was transferred to a QIAamp® Mini spin column and underwent the manufacturer’s standard protocol from the QIAamp® DNA Blood Mini Kit for column washing. DNA was eluted in pre-warmed (56°C) sterile water twice in 50 μL volumes. To assess DNA yield, a qPCR assay targeting the β-globin gene against a standard curve was performed, and only samples anticipated to contain at least 1,000 T cells were sent for TCRβ sequencing. Maternal PBMC underwent genomic DNA extraction using the standard protocol provided in the QIAamp® DNA Blood Mini Kit (Qiagen), and 3.4 μg of total genomic DNA was sent for TCRβ sequencing. Samples were sent in batches to Adaptive Biotechnologies (Seattle, WA, USA) and assayed using their ImmunoSEQ® hsTCRBB (*46*) service pipeline. Sequencing was performed at a survey level following Adaptive Biotechnologies’ custom protocol. Quality control of sequencing data was performed by Adaptive Biotechnologies. Total productive templates from PBMC ranged from 64,403 to 118,002 (median 94,389), whereas total productive templates from BMC ranged from 936 to 17,085 (median 2,976) (File S1).

### TCR sequence analysis

TCRβ sequence data were analyzed using the immunoSEQ® Analyzer software (Adaptive Biotechnologies) and/or exported to R for analysis with the package immunoArch (*47*) or divo (*48*). Repertoire overlap between blood and breastmilk was assessed using the Morisita index (immunoSEQ Analyzer®) and the power geometric index (divo), which are relatively protected from differences in sampling depth (*29*). To identify overabundant clones in the breastmilk, frequencies of TCR clonotype nucleotide sequences in the breastmilk were compared to those in peripheral blood using the Differential Abundance tool in immunoSEQ® Analyzer using the binomial statistical method with Benjamini Hochberg correction and a lower limit of detection of 10.

The CDR3β amino acid sequence of every overabundant clone was compared to that of all other overabundant clones within each participant using tcrdist3 (*31, 32*). Similarly, CDR3β amino acid sequences of overabundant clones across all participants were compared to one another using tcrdist3 (*31, 32*). To identify epitope specificity of overabundant breastmilk clones, TCRβ sequences were matched by CDR3 amino acid and V gene identification against several public databases of TCRβ epitope specificity, namely ImmuneCODE (*36*) (available online at https://clients.adaptivebiotech.com/pub/covid-2020), VDJdb (available at https://vdjdb.cdr3.net) (*49*), TCRBdb (available online at http://bioinfo.life.hust.edu.cn/TCRdb/#/) (*50*), McPAS-TCRB (available online at http://friedmanlab.weizmann.ac.il/McPAS-TCRB/) (*51*), and Immune Epitope Database and analysis Resource (IEDB) (available online at http://www.iedb.org/home_v3.php) (*52*). Epitope matches from any of these databases were considered a direct match if the CDR3 amino acid and TRBV gene were identical and were considered a predicted match if the CDR3 amino acid sequences were identical, but V gene usage was mismatched. All overabundant breastmilk clones were also queried using the IEDB TCRMatch Tool (available online at http://tools.iedb.org/tcrmatch/) (*35*) with a score threshold of 0.97 to identify closely related epitope restrictions.

TCRβ sequences from both breastmilk and PBMC were evaluated for candidate Spike-specific restriction using the COVID Search Tool in immunoSEQ® Analyzer, which utilizes TCRβ sequences assigned as specific for SARS-CoV-2 from the ImmuneCODE database (*36, 42*). We compared the frequency of Spike-specific clones out of the number of non-Spike SARS2-specific clones in each sample, where non-Spike SARS2-specific clones were used to represent the “background” response noted in PBMC to identify a vaccine-specific response. Frequencies of assigned Spike-specific TCRβ sequences were compared with negative binomial models accounting for the total productive templates in each sample. For comparison of clonality metrics only (Simpson Clonality and maximum productive frequency), the full dataset of paired breastmilk and PBMC TCRβ sequences was down-sampled to the lowest productive template frequency, and metrics were calculated and compared using the down-sampled dataset.

Candidate spike-specific TCRβ in breastmilk and PBMC were further validated by comparing CDR3β amino acid sequences, V gene usage, epitope HLA restriction, and participant HLA concordance to an internally vetted dataset of published TCRβ sequences obtained via Spike - epitope-loaded tetramer or multimer-based experiments (*37-41*). An exact match at the TCRβ CDR3 sequence was required for these analyses. The internally vetted dataset (*37-41*) was used to train the tcrdist3 algorithm (*31, 32*) with a distance unit threshold of 10 to identify additional potential Spike-specific TCRβ in breastmilk.

### ELISA

Wells of a sterile high-binding 384-well plate (Corning) were coated with 11.8 μM full-length trimeric SARS-CoV-2 spike protein (Institute for Protein Design, University of Washington) or 1 μg/mL RBD (Institute for Protein Design, University of Washington) for at least 2 hours at room temperature or up to 3 days at 4°C. Each well was washed three times with 100 μL 1X Wash Buffer A (Teknova) using a BioTek EL406 plate washer (BioTek). Wells were blocked with with 100 μL blocking buffer (PBS, 1% BSA w/v, 0.05% Tween-20) at least two hours at room temperature or at 4 °C overnight. Wells were then washed three times with 1X Wash Buffer A, and 50 μL assay diluent (50% 1X Wash Buffer A, 50% PBS, 0.1% BSA) was added to each well. Aqueous breast milk fractions were diluted 1:2 and plasma samples were diluted 1:20 in assay diluent in a separate 96-well plate and then added to the first column of the 384-well plate, further diluting the sample by a factor of 5. Each sample was then serially diluted 1:5 in wells from left to right across the plate. After incubating overnight at 4 °C, plates were brought to room temperature, and wells were rinsed five times with 1X Wash Buffer A. HRP-conjugated immunoglobulins were diluted in assay diluent as shown in Table S8, and then 50 μL was added to the appropriate wells. Wells incubated with secondary antibodies for 1 hour protected from light at room temperature and were washed five times with 1X Wash Buffer A followed by one wash with PBS. Wells were then treated with 50 μL Tetramethylbenzidine (TMB, SeraCare) for 2 to 10 minutes, depending on the secondary antibody (Table S8), when the reaction was stopped with the addition of 25 μL 1N H_2_SO_4_ (Millipore Sigma). Plates were read at 450 nm with a reference filter set at 570 nm using a SpectraMax 13x plate reader (Molecular Devices) and SoftMax Pro 6.4.2 analysis software. The average optical density (OD) value for all dilutions of the negative sample was used to set a minimum cutoff value for each plate. Plate cutoff values were then used to calculate each sample’s end point titer (EPT) using a 4-parameter logistic model in XL-fit software (model 208, IDBS). The EPT value for each duplicate plate was averaged for a final EPT run value for each sample.

### Statistical approach

To assess the primary difference between cellular phenotypic frequency (e.g. % CCR9+ of CD4+ T cells) in BMC versus PBMC, we built a linear regression model for each outcome with sample type as the predictor, adjustment for time since delivery, and clustering by individual to account for the correlation of individuals contributing paired BMC and PBMC. Due to the large difference in sampling depth in the two compartments, negative binomial models were used to compare the frequency of Spike-specific templates in BMC versus PBMC, accounting for both the number of total SARS2-specific templates identified (viral genome wide) and the number of Spike -specific templates identified to determine the enrichment of Spike above background reactivity to non - SARS2 coronaviruses and/or TCRs present in the naïve repertoire, and adjusting for time since delivery. The negative binomial model generates an incidence rate ratio (IRR) which represents the number of Spike-specific templates found in the experimental group (e.g. BMC) for every one template identified in the control group (e.g. PBMC). The phenotype and frequency of tetramer positive T cells in the BMC of individuals pre- and post-3^rd^ dose was compared with paired t tests. A p-value less than 0.05 was considered significant.

## Data Availability

All data produced in the present study are contained in the  manuscript or are available upon reasonable request to the authors.

## Supplementary Materials

Materials and Methods

Figs. S1 to S8

Tables S1 to S8

## Author contributions

Conceptualization: BA, YJ, WEH

Methodology: YJ, MC, ESF, SJ, JH, XW, LJ, TP, SEL, RNC, DMK, WEH

Participant enrollment: AK, WY, TN, WEH

Investigation: BA, YJ, MC, ESF, SJ, JH, XW, LJ, TP, DMK, WEH

Visualization: BA, MC, WEH

Funding acquisition: BA, ESF, SJ, AK, RNC, DMK, WEH Supervision: NM, RNC, DMK, WEH

Writing –draft: BA, YJ, WEH

Writing – critical review/editing: BA, YJ, ESF, SJ, XW, NM, SEL, DMK, WEH

## Acknowledgements

We thank all those who donated blood and breastmilk for this work as well as Drs. Christine Johnston and Anna Wald for providing control specimens and the NIH Tetramer Core Facility for synthesizing the HLA-A*01_LTD-APC and HLA-A*24_NYN-APC tetramers.

## Funding

NIH grant T32AI007509 (BA)

NIH grant K08AI148588 (ESF)

NIH grant T32CA080416 (SJ)

NIH grant K23AI153390-01 (AK)

NIH grant 1UM1AI148373-01 (RNC)

NIH grant 3UM1 AI148373-01S1 (RNC)

NIH grant R21AI163999 (DMK)

NIH grant K08AI135072 (WEH)

NIH contract 75N93019C00063 (DMK)

Burroughs Wellcome Fund CAMS grant 1017213 (WEH)

University of Washington and Seattle Children’s (WEH)

## Data and materials availability

All flow cytometry data are available in the main manuscript or supplementary files, and all TCR sequencing data will be deposited in immuneACCESS®.

