## Supplementary Material for "Spike-specific T cells are enriched in breastmilk following SARS-CoV-2 mRNA vaccination"

Figs. S1 to S8

Tables S1 to S8

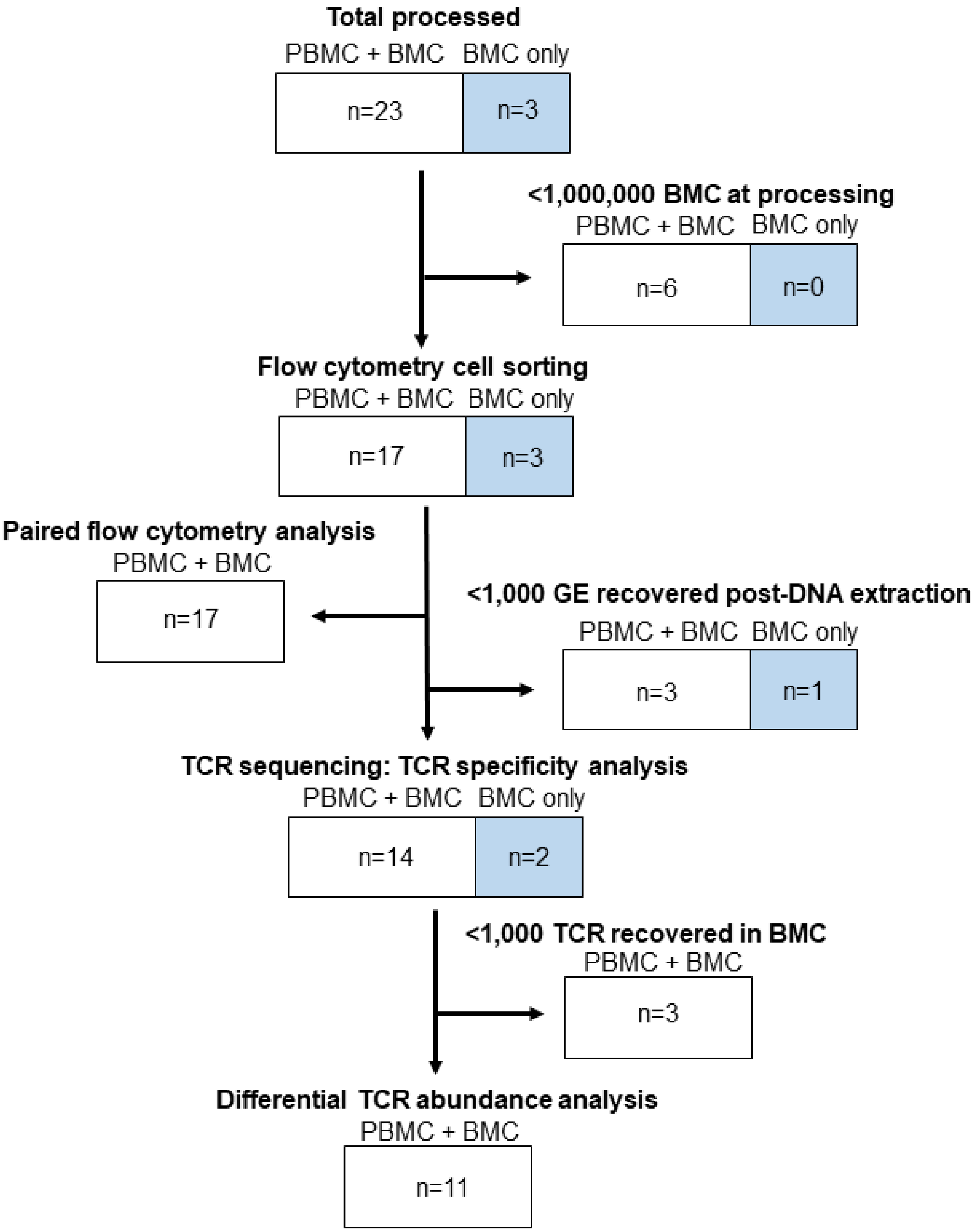


**Fig. S1. Inclusion criteria for study analyses.** Breastmilk was collected from n=26 individuals, n=23 of which also contributed blood samples. Breastmilk and blood were processed for breastmilk cells (BMC) and peripheral blood mononuclear cells (PBMC), respectively. Individuals (n=6) were excluded from study analyses if <1,000,000 BMC were recovered at processing. Flow cytometry cell sorting was performed for n=20 individuals. Paired flow cytometry analysis was performed on samples for which both BMC and PBMC were available and at least 100 CD4+ and

CD8+ T cells were recorded (n=17). If <1,000 DNA genomic equivalents (GE) were recovered from sorted BMC, individuals (n=4) were excluded from TCR sequencing and TCR specificity analysis (n=16 included). If TCR sequencing yielded fewer than 1,000 TCR templates, individuals (n=3) were excluded from differential TCR abundance analysis. Individuals which met the above criteria and had both BMC and PBMC available (n=11) were included in differential TCR abundance analysis. Spike tetramer staining on pre- and post-3rd SARS2 mRNA vaccine dose samples was performed for n=5 individuals who returned from the original cohort and n=3 newly enrolled individuals.

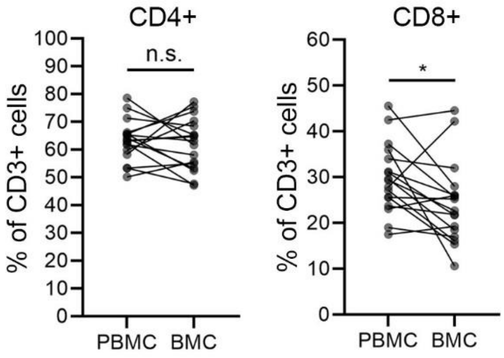


**Fig. S2. CD4+ and CD8+ T cell relative frequency in breastmilk and peripheral blood.** Frequencies of CD4+ and CD8+ among CD3+ cells in BMC and PBMC were determined by flow cytometry (n=17). Comparisons made with linear regression and clustering by individual, $^*p < 0.05$, n.s. (not significant). CD4+ (PBMC vs. BMC): 63% vs. 62%, p=0.6; CD8+ (PBMC vs. BMC): 30% vs. 24%, p=0.03.

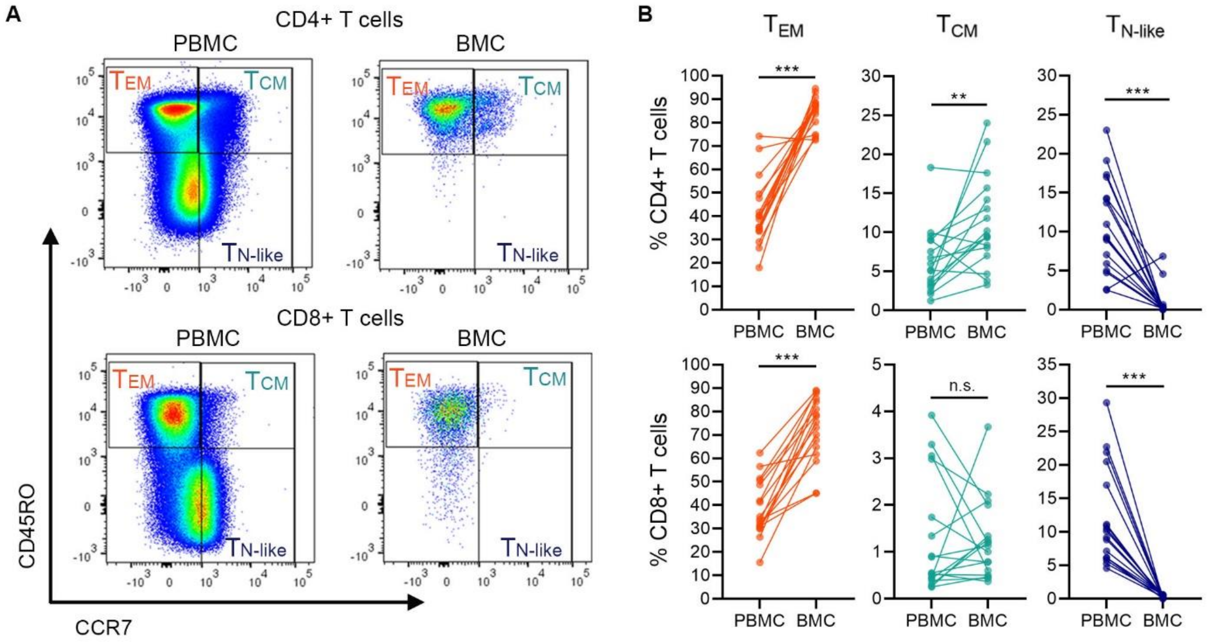


**Fig. S3. Antigen-experienced T cells are enriched in breastmilk.** The distribution of memory and naïve populations of CD4+ and CD8+ T cells within PBMC and BMC (n=17). Comparisons made with linear regression and clustering by individual, ***p< 0.001, **p< 0.01, *p< 0.05, n.s. (not significant). (**A**) Scatter plots from one representative participant. (**B**) Frequencies of $T_{EM}$, $T_{CM}$, and $T_{N\text{-like}}$ CD4+ and CD8+ T cells in PBMC and BMC: CD4+ $T_{EM}$: 41% vs. 84%, p<0.001, CD4+ $T_{CM}$: 6% vs. 11%, p=0.006, CD4+ $T_{N\text{-like}}$: 11% vs. 1%, p<0.001; CD8+ $T_{EM}$: 38% vs.73%, p< 0.001, CD8+ $T_{CM}$: 1% vs. 1%, p=n.s., CD8+ $T_{N\text{-like}}$: 12% vs. 0.2%, p<0.001.

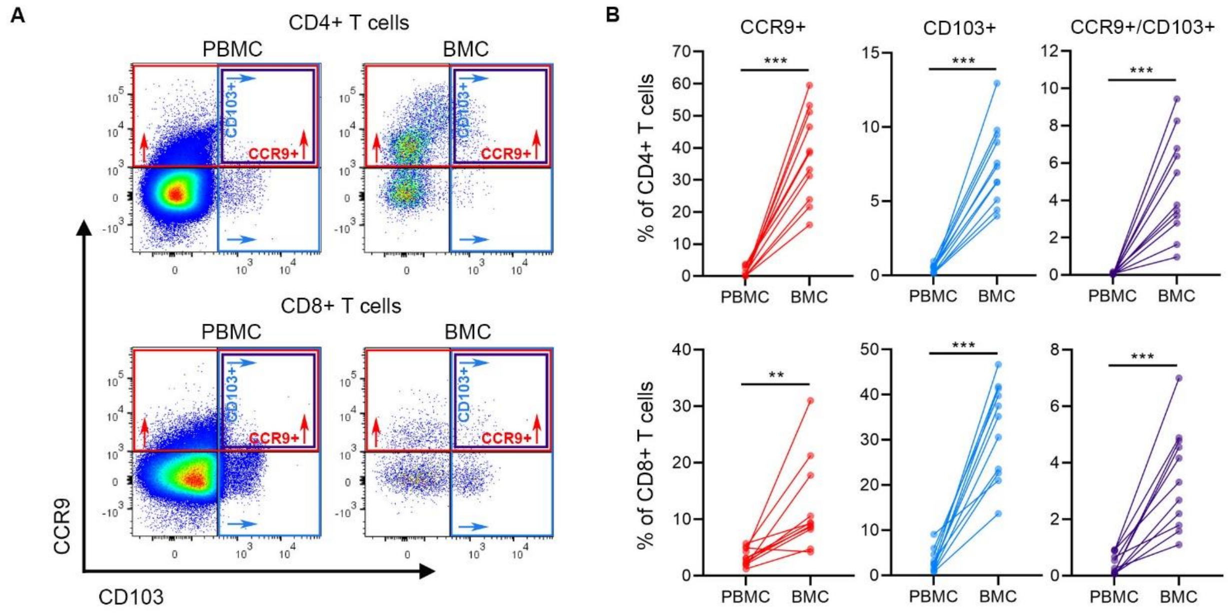


**Fig. S4**. **Breastmilk T cells display high levels of mucosal-homing markers.** CCR9 and CD103 expression in CD4+ and CD8+ T cell populations from PBMC and BMC (n=11). Comparisons made with linear regression and clustering by individual, ***p<0.001, **p<0.01, *p<0.05. (**A**) Scatter plots from one representative participant. (**B**) Expression of CCR9 and CD103 within CD4+ and CD8+ T cell populations in PBMC and BMC: CD4+/CCR9+: 4% vs. 38%, p<0.001, CD4+/CD103+: 0.4% vs. 7%, p <0.001, CD4+/CCR9+/CD103+: 0.1 vs. 5%, p<0.001, CD8+/CCR9+: 3% vs. 12%, p=0.005; CD8+/CD103+: 3% vs. 32%, p<0.001, CD8+/CCR9+/CD103+: 0.4% vs. 3%, p<0.001.

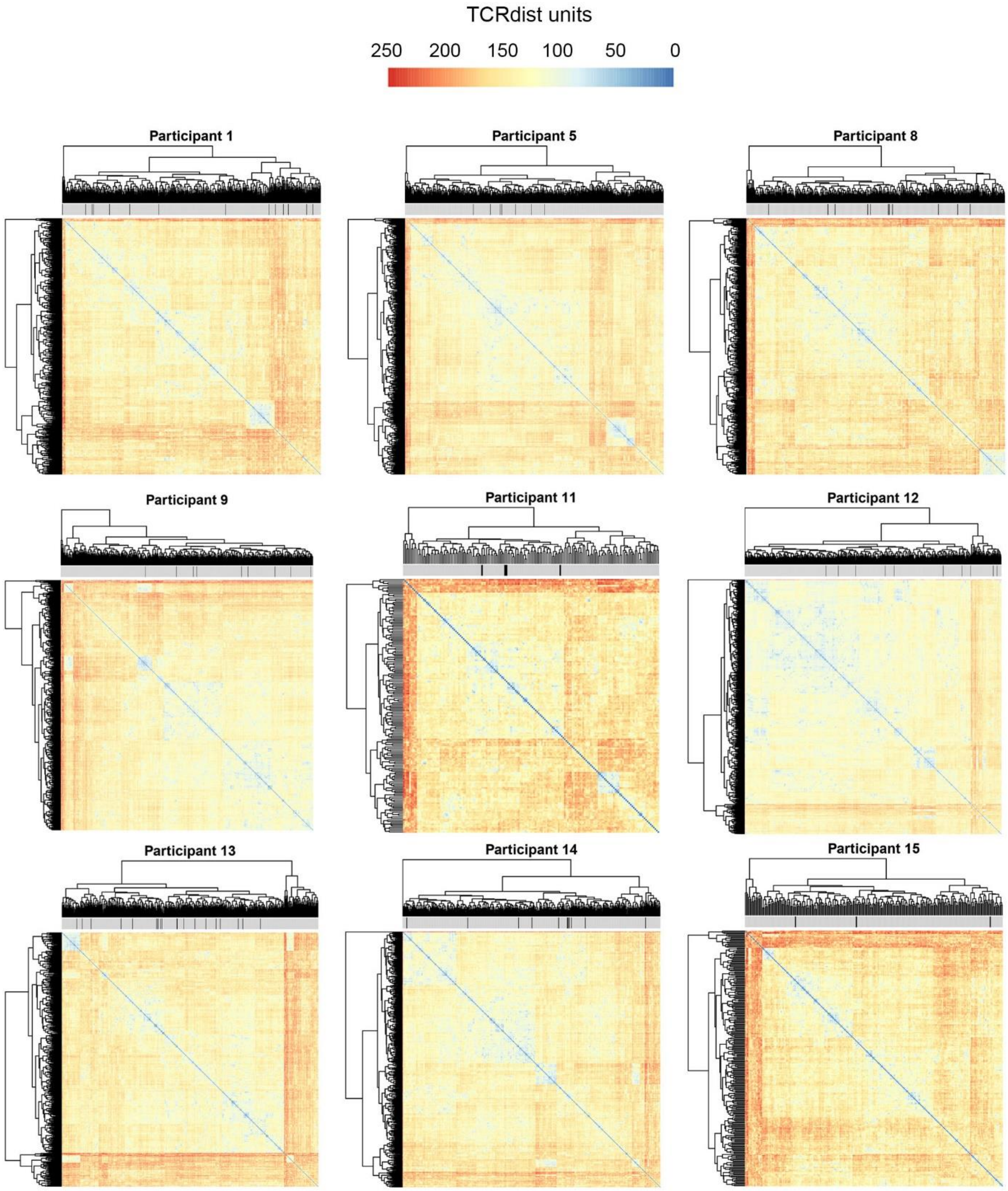


**Fig. S5. Overabundant TCR clones in breastmilk are diverse.** Each TCRβ CDR3 amino acid sequence obtained from a participant's BMC was compared to all other TCRβ CDR3 amino acid sequences within that participant's BMC using tcrdist3 (n=11, n=9 shown). Heatmaps depicting distance units and clustering within each participant's breastmilk T cell repertoire are shown.

Black ticks denote TCRβ sequences that were significantly enriched in BMC relative to PBMC in this participant. Data from n=2 participants shown in Fig. 2A.

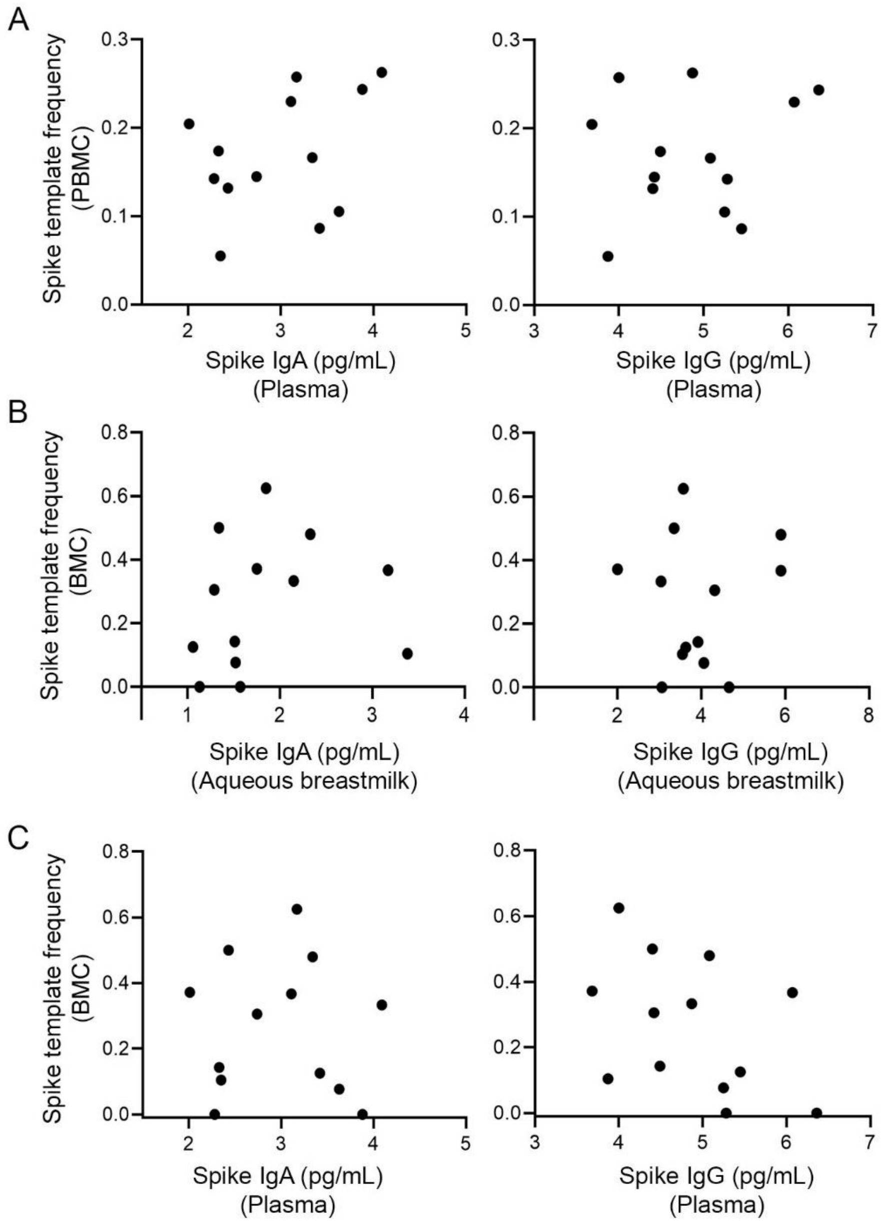


**Fig. S6. Spike-specific antibody levels and the frequency of spike-specific T cells were not strongly correlated.** Data were analyzed using a negative binomial regression model adjusting for time since delivery (n=13). (**A**) The frequency of Spike-predicted TCRβ templates in PBMC are shown as a function of Spike-specific IgA level (left) and IgG level (right) in the plasma. Spike-specific IgA by Spike-specific TCR frequency: IRR=1.1, 95% C.I. (0.85-1.5), p=0.4; Spike-specific IgG by spike-specific TCR frequency: IRR=1.1, 95% C.I. (0.90-1.3), p=0.4 (**B**) The frequency of Spike-predicted TCRβ templates in BMC are shown as a function of Spike-specific IgA level (left) and IgG level (right) in aqueous breastmilk. Spike-specific IgA by Spike-specific TCR frequency: IRR=0.95, 95% C.I. (0.66-1.4), p=0.8; spike-specific IgG by spike-specific TCR frequency:

IRR=0.97, 95% C.I. (0.88-1.1), p=0.6. (**C**) The frequency of spike-predicted TCRβ templates in BMC are shown as a function of Spike-specific IgA level (left) and IgG level (right) in the plasma. Spike-specific IgA by Spike-specific TCR frequency: IRR=0.86, 95% C.I. (0.59-1.3), p=0.4; spike-specific IgG by spike-specific TCR frequency: IRR=0.9, 95% C.I. (0.68-1.1), p=0.3.

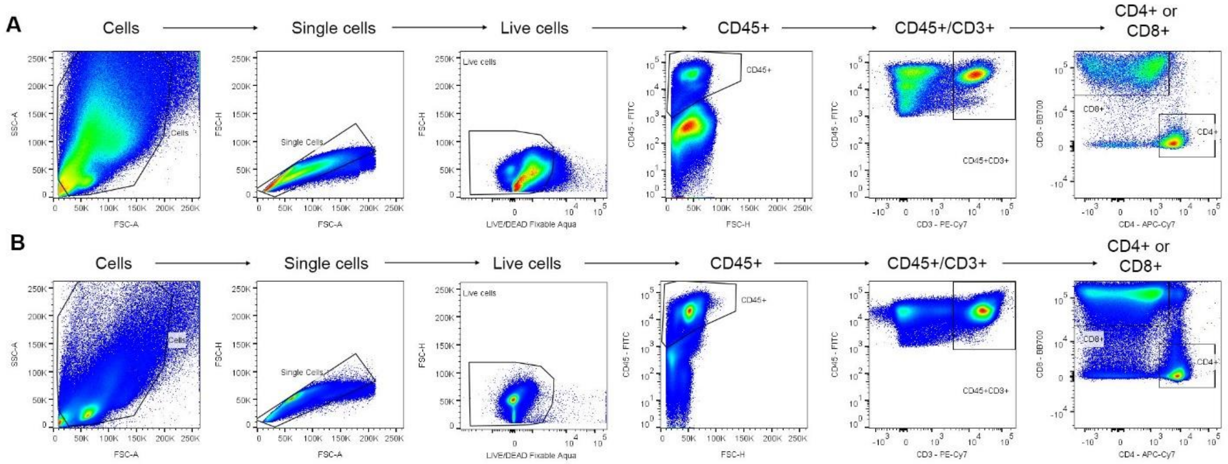


**Fig. S7. Gating strategy for phenotyping BMC and PBMC.** The gating strategy used to analyze (**A**) BMC and (**B**) PBMC that were stained with flow cytometry panel 1 is shown.

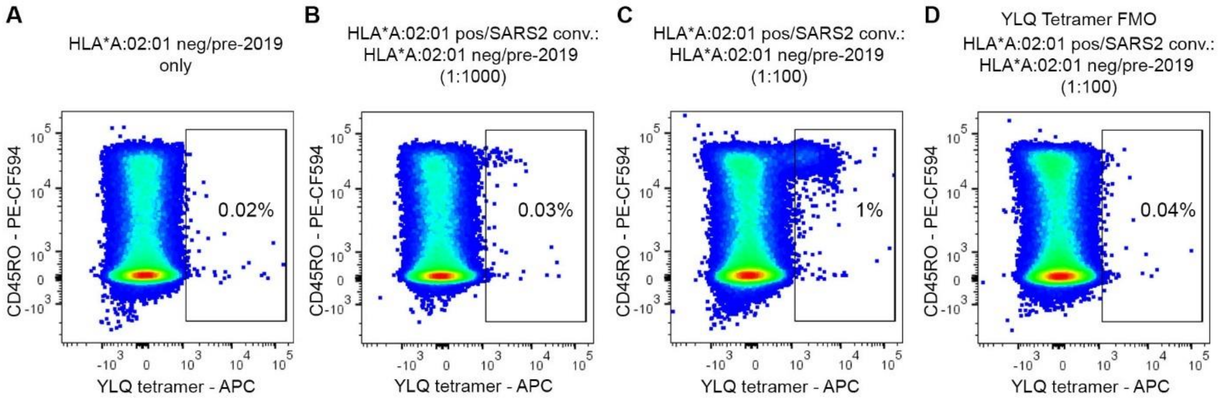


**Fig. S8. Example validation of Spike-loaded tetramer HLA*A02_YLQ.** Positive and negative controls were analyzed for HLA-A*02:01-YLQ tetramer positive cells. (**A**) PBMC from an HLA-A*02:01 negative donor obtained prior to 2019. (**B**) PBMC from an HLA-A*02:01 positive, SARS2 convalescent donor were mixed with PBMC from an HLA-A*02:01 negative donor obtained prior to 2019 at a ratio of 1:1000. (**C**) PBMC from an HLA-A*02:01 positive, SARS2 convalescent donor were mixed with PBMC from an HLA-A*02:01 negative donor obtained prior to 2019 at a ratio of 1:100. (**D**) The fluorescence minus one (FMO) HLA-A*02:01-YLQ tetramer control for the PBMC mixture described in (**C**) is shown.

| Participant | Templates | CDR3 amino acid sequence | Restriction | Match | PMID of matching sequence (if available) |
|---|---|---|---|---|---|
| 10 | 120 | CASSDYEQYF | M. tuberculosis lysate | Direct | 32341563 |
| 13 | 27 | CASSIGLYGYTF | Influenza M1/SARS-2 Spike | Direct | 25609818, 28423320, 32793919 |
| 13 | 6 | CASSSRSSYEQYF | Influenza M1/SARS-2 Spike | Direct | 28423320, 32793919 |
| 13 | 57 | CASSPGQGEGYEQYF | EBV BZLF1 | Direct | 1X LIT000047 RevC Application Note: A new way of exploring immunity https://pages.10xgenomics.com/rs/446-PBO-704/images/10x_AN047_IP_A_New_Way_of_Exploring_Immunity_Digital.pdf |
| 16 | 10 | CASSLNRGPSDTQYF | Influenza M1 | Direct | 28636589 |
| 5 | 19 | CASSLMAYEQYF | SARS-2 Orf1ab | Predicted | 32793919 |
| 9 | 6 | CASSDGYSNQPQHF | SARS-2 Spike/SARS-2 Membrane protein | Predicted | 32793919, 32793896 |
| 10 | 264 | CASSPGTGGEQYF | SARS-2 Orf3a | Predicted | 32793919 |
| 10 | 56 | CASSQDRETQYF | SARS-2 Orf7b | Predicted | 32793919 |
| 10 | 25 | CASRPGQGNNEQFF | SARS-2 Spike | Predicted | 32793919 |
| 10 | 11 | CASSHPGQGTYEQYF | EBV IE63 homolog | Predicted | 32184241 |
| 10 | 71 | CASSLGTGTYEQYF | SARS-2 Spike | Predicted | 32793896 |
| 10 | 39 | CASSFPNEQFF | SARS-2 Spike | Predicted | 32793896 |
| 14 | 12 | CASSPNNEQFF | M. tuberculosis lysate/SARS-2 Membrane protein | Predicted | 32341563, 32793919 |

| | | | | | |
|---|---|---|---|---|---|
| 14 | 11 | CASSLLGGQPQHF | SARS-2 Orf1ab | Predicted | 32793919 |
| 16 | 10 | CASSSPGTGVNEQFF | SARS-2 Spike/SARS-2 Orf7a | Predicted | 32793919 |

**Table S1. Potential antigen specificities of overabundant breastmilk clones identified by CDR3 amino acid sequence comparison to TCR sequence databases McPAS-TCR, VDJdb, TCRdb, ImmuneCODE, and Immune Epitope Database and analysis Resource (IEDB) (also shown in Fig. 4).** A direct match indicates that the CDR3 amino acid sequence and V gene usage of the overabundant clone was identical to published reports. A predicted match indicates that the CDR3 amino acid sequence was identical to published reports, but V gene usage was not identical.

| Participant | CDR3b amino acid sequence of overabundant breastmilk clone | Templates | Productive frequency | CDR3b amino acid sequence(s) of published match | Cognate antigen origin | Cognate antigen | Amino acid sequence of cognate antigen epitope |
|---|---|---|---|---|---|---|---|
| 5 | CASSLMAYEQYF | 19 | 0.01710171 | ASSLVAYEQY | SARS-2 | Orf10 | MGYINVFAFPFTIYSL |
| | | | | ASSLVAYEQY | SARS-2 | Orf7b | MIELSLIDFYLCFLAFLLFLVLIML |
| | | | | ASSLMSYEQY | SARS-2 | Orf1ab | FLPRVFSAV |
| 5 | CASSLSGVEQYF | 12 | 0.01080108 | ASSLAGVEQY | SARS-2 | Nuc | LSPRWYFYYL |
| | | | | ASSLSGLEQY | SARS-2 | Orf3a | NPLLYDANYFLCW |
| | | | | ASSLAGIEQY | SARS-2 | Nuc | LSPRWYFYYL |
| | | | | ASSLAGVEQF | SARS-2 | Nuc | LSPRWYFYYL |
| 8 | CASSIGTGDQPQHF | 11 | 0.003696237 | ASSLGTGDQPQH | SARS-2 | Nucleocapsid | GMEVTPSGTWLTY |
| 9 | CASSDGYSNQPQHF | 6 | 0.006410256 | ASSEGYSNQPQH | SARS-2 | Orf1ab | HTTDPSFLGRY |
| | | | | ASSEGYSNQPQH | SARS-2 | Orf1ab | LLLDDFVEII |
| 9 | CASSVGGTGNTIYF | 6 | 0.006410256 | ASSLGGTGNTIY | SARS-2 | Orf3a | GLEAPFLYLYALVYFLQSINFVRIIMR |
| 10 | CASSLGQDTEAFF | 165 | 0.009657594 | ASSLGQETEAF | SARS-2 | Orf7a | VQELYSPIFLIV |
| 10 | CASGIGTEAFF | 158 | 0.009247878 | ASGLGTEAF | SARS-2 | Orf10 | MGYINVFAFPFTIYSL |
| 10 | CASSLGTGTYEQYF | 71 | 0.004155692 | ASSVGTGTYEQY | SARS-2 | Nucleocapsid | GMEVTPSGTWLTY |
| 10 | CASSLKVTDTQYF | 56 | 0.003277729 | ASSLKVSDTQY | SARS-2 | Orf1ab | VPHVGEIPVAYRKVLL |
| | | | | ASSLKATDTQY | SARS-2 | Orf1ab | SNEKQEILGTVSWNL |
| 10 | CASSQDRETQYF | 56 | 0.003277729 | ASSQDRDTQY | CMV | glycoprotein | VMAPRTLIL |
| | | | | ASSQDRKTQY | SARS-2 | Orf7b | MIELSLIDFYLCFLAFLLFLVLIML |
| 10 | CASSSSYSNQPQHF | 35 | 0.002048581 | ASSASYSNQPQH | SARS-2 | Spike precursor | FIAGLIAIV |
| | | | | ASSSYSNQPQH | SARS-2 | Nucleocapsid | AQFAPSASAFFGMSR |
| 10 | CASSTGQGDYNEQFF | 35 | 0.002048581 | ASSSGQGDYNEQF | Hepatitis B virus | Core/pre core | RTQSPRRR |
| 10 | CASSIGGQQYF | 16 | 9.36E-04 | ASSIGGEQY | Influenza | PBP2 | SSLENFRAYV |

| | | | | | | | |
|---|---|---|---|---|---|---|---|
| | | | | ASSLGGQQF | CMV | immediate early protein 1 | KLGGALQAK |
| 10 | CASSIVAGGSTDTQYF | 13 | 7.61E-04 | ASSVLAGGSTDTQY | EBV | NA4 | AVFDRKSDAK |
| 10 | CASSYMAGAYEQYF | 10 | 5.85E-04 | ASSYLAGAYEQY | SARS-2 | Orf1ab | QLMCQPILLL |
| 10 | CASSRDTGELFF | 9 | 5.27E-04 | ARSRDTGELF | EBV | BRLF1 | YVLDHLIVV |
| 13 | CASSIGLYGYTF | 27 | 0.007058824 | ASSIGIYGYT | Influenza | Matrix | GILGFVFTL |
| | | | | ASSLGLYGYT | Influenza | Matrix | GILGFVFTL |
| | | | | ASSIGVYGYT | Influenza | Matrix | GILGFVFTL |
| | | | | ASSVGIYGYT | Influenza | Matrix | GILGFVFTL |
| | | | | ASSMGLYGYT | Influenza | Matrix | GILGFVFTL |
| | | | | ASSLGIYGYT | Yellow Fever Virus | NS4b | LLWNGPMAV |
| | | | | ASSLGIYGYT | Hepatitis B virus | DNA polymerase | LVVDFSQFSR |
| 13 | CASSSRSSYEQYF | 6 | 0.001568627 | ASSTRSSYEQY | Influenza | Matrix | GILGFVFTL |
| | | | | ASSTRSSYEQY | EBV | IE63 homolog | GLCTLVAML |
| | | | | AISSRSSYEQY | CMV | UL83 | NLVPMVATV |
| | | | | ASSRSSYEQY | Influenza | Matrix | GILGFVFTL |
| | | | | ASSRSSYEQY | SARS-2 | Orf3a | FVCNLLLLFVTVYSHLLLV |
| | | | | ASSSRSSYDQY | Influenza | Matrix | GILGFVFTL |
| | | | | ASSSRTSYEQY | Influenza | Matrix | GILGFVFTL |
| | | | | ASSSRSAYEQY | Influenza | Matrix | GILGFVFTL |
| | | | | ASSSRASYEQY | Influenza | Matrix | GILGFVFTL |
| | | | | ASSARSSYEQY | Influenza | Matrix | GILGFVFTL |

| | | | | | | | |
|---|---|---|---|---|---|---|---|
| | | | | ASSARSSYEQY | EBV | IE63 homolog | RAKFKQLL |
| | | | | ASSARSSYEQY | EBV | IE63 homolog | GLCTLVAML |
| | | | | ASSQRSSYEQY | Influenza | Matrix | GILGFVFTL |
| | | | | ASSNRSSYEQY | Influenza | Matrix | GILGFVFTL |
| | | | | ASSKRSSYEQY | Influenza | Matrix | GILGFVFTL |
| 14 | CASSPNNEQFF | 12 | 0.003183024 | ASTPNNEQF | SARS-2 | Orf7b | MIELSLIDFYLCFLAFLLFLVLIML |
| 14 | CASSLLGGQPQHF | 11 | 0.002917772 | ASSLMGGQPQH | SARS-2 | Orf1ab | KLSYGIATV |
| 16 | CASSLNRGPSDTQYF | 10 | 0.002044572 | ASSLNRGPADTQY | Influenza | Matrix | GILGFVFTL |
| 16 | CASSPLTGSSYEQYF | 6 | 0.001226743 | ASSPLTGASYEQY | SARS-2 | Spike | KLPDDFTGCV |
| 16 | CASNQGGSYEQYF | 4 | 8.18E-04 | ASSQGGSYEQY | SARS-2 | Orf1ab | LVLSVNPYV |
| | | | | ASSQGGSYEQY | SARS-2 | Membrane protein | SYFIASFRLFA |

**Table S2. Predicted epitope specificities of overabundant breastmilk clones identified by the IEDB TCRMatch Tool.**

| Participant | Breastmilk-derived CDR3b amino acid sequence | V gene of reference sequence | V gene of breastmilk derived TCR (this study) | Cognate Spike-specific epitope (reference) | HLA restriction (reference) | Relevant HLA allele of participant (this study) | Templates (this study) | Productive frequency (this study) | Reference PMID |
|---|---|---|---|---|---|---|---|---|---|
| 1 | CASSLAGYEQYF | TRBV5-1 | TCRBV11-02*01 | LTDEMIAQY | A*01:01 | A*01:01:01 | 1 | 0.00022 | 34341799 |
| 1 | CASSPGQGYEQYF | TRBV6-4 | TCRBV27-01*01 | NYNYLYRLF | A*24:02 | None | 1 | 0.00022 | 34341799 |
| 2 | CASGEENTGELFF | TRBV5-1 | TCRBV05-01*01 | YLQPRTFLL | A*02:01 | A*02:01:01 | 3 | 0.01415 | 33326767 |
| 2 | CASSPPGGGNTGELFF | TRBV18 | TCRBV05-08*01 | YLQPRTFLL | A*02:01 | A*02:01:01 | 3 | 0.01415 | 33326767 |
| 3 | CASSPDIEQYF | TRBV7-9 | TCRBV07-09*01 | YLQPRTFLL | A*02:01 | Typing not available | 3 | 0.00095 | 34341799 |
| | | TRBV7-9 | | YLQPRTFLL | A*02:01 | Typing not available | | | 33326767 |
| | | TRBV7-9 | | YLQPRTFLL | A*02:01 | Typing not available | | | 33951417 |
| 3 | CASSLQNTGELFF | 7-8 | TCRBV07-08*03 | YLQPRTFLL | A*02:01 | Typing not available | 1 | 0.00032 | 33951417 |
| 3 | CASSLDIEAFF | TRBV7-9 | TCRBV07-09*01 | YLQPRTFLL | A*02:01 | Typing not available | 10 | 0.00318 | 34341799 |
| | | TRBV7-9 | | YLQPRTFLL | A*02:01 | Typing not available | | | 33326767 |
| | | TRBV7-9 | | YLQPRTFLL | A*02:01 | Typing not available | | | 33951417 |
| 3 | CASSDGNTGELFF | TRBV5-1 | TCRBV05-01*01 | YLQPRTFLL | A*02:01 | Typing not available | 1 | 0.00032 | 33326767 |
| 3 | CASSDTNTGELFF | TRBV5-1 | TCRBV02-01 | YLQPRTFLL | A*02:01 | Typing not available | 2 | 0.00064 | 33326767 |

| | | | | | | | | | |
|---|---|---|---|---|---|---|---|---|---|
| | | TRBV7-9 | | YLQPRTFLL | A*02:01 | Typing not available | | | 33326767 |
| 3 | CASSPDIEAFF | TRBV7-9 | TCRBV07-09*01 | YLQPRTFLL | A*02:01 | Typing not available | 7 | 0.00223 | 34341799 |
| | | TRBV7-9 | | YLQPRTFLL | A*02:01 | Typing not available | | | 33951417 |
| 6 | CASQDTNTGELFF | TRBV2 | TCRBV06-04*01 | YLQPRTFLL | A*02:01 | Typing not available | 1 | 0.00029 | 33951417 |
| 6 | CASSLAGSSYNSPLHF | TRBV7-8 | TCRBV05-01*01 | YLQPRTFLL | A*02:01 | Typing not available | 1 | 0.00029 | 34341799 |
| 6 | CASSLVAGGNTGELFF | TRBV13 | TCRBV06-04*01 | YLQPRTFLL | A*02:01 | Typing not available | 1 | 0.00029 | 33326767 |
| 6 | CASGGTNTGELFF | TRBV2 | TCRBV07-03*01 | YLQPRTFLL | A*02:01 | Typing not available | 1 | 0.00029 | 34341799 |
| | | TRBV2 | | YLQPRTFLL | A*02:01 | Typing not available | | | 33326767 |
| 6 | CASSLAGYEQYF | TRBV5-1 | TCRBV12-03/12-04*01 | LTDEMIAQY | A*01:01 | Typing not available | 1 | 0.00029 | 34341799 |
| 8 | CASSPGQGYEQYF | TRBV6-4 | TCRBV18-01*01 | NYNYLYRLF | A*24:02 | None | 2 | 0.00067 | 34341799 |
| | | TRBV2 | | YLQPRTFLL | A*02:01 | A*02:01:01 | | | 34341799 |
| 8 | CASSDLDTGELFF | TRBV2 | TCRBV02-01 | YLQPRTFLL | A*02:01 | A*02:01:01 | 1 | 0.00034 | 33326767 |
| | | TRBV2 | | YLQPRTFLL | A*02:01 | A*02:01:01 | | | 33951417 |
| 8 | CASSPLAGGNTGELFF | TRBV3-1 | TCRBV05-04*01 | YLQPRTFLL | A*02:01 | A*02:01:01 | 1 | 0.00034 | 33326767 |
| 10 | CASSLQNTGELFF | 7-8 | TCRBV07-09*01 | YLQPRTFLL | A*02:01 | None | 1 | 0.00006 | 33951417 |
| 10 | CASSGGNTIYF | TRBV28 | TCRBV11-03*01 | LTDEMIAQY | A*01:01 | None | 3 | 0.00018 | 34341799 |
| 10 | CASSLAGYEQYF | TRBV5-1 | TCRBV07 | LTDEMIAQY | A*01:01 | None | 1 | 0.00006 | 34341799 |
| 10 | CASSLAGYEQYF | | TCRBV05-05*01 | | | None | 1 | 0.00006 | |

| | | | | | | | | | |
|---|---|---|---|---|---|---|---|---|---|
| 14 | CASSLGGNQPQHF | TRBV12-3 | TCRBV05-01*01 | LTDEMIAQY | A*01:01 | None | 1 | 0.00027 | 34341799 |
| 16 | CASSGTSGSTDTQYF | TRBV6-4 | TCRBV06-04*01 | NQKLIANQF | B*15:01 | B*15:01:01 | 1 | 0.00020 | 34341799 |
| 16 | CASSLEDTNYGYTF | TRBV7-2 | TCRBV07-02 | NQKLIANQF | B*15:01 | B*15:01:01 | 2 | 0.00041 | 34341799 |
| 16 | CASSQGGNEQYF | TRBV4-1 | TCRBV03-01/03-02*01 | YLQPRTFLL | A*2:01 | A*02:01:01 | 2 | 0.00041 | 34341799 |

**Table S3. Breastmilk-derived TCR sequences that match previously published sequences obtained from SARS2 Spike-loaded tetramer or multimer experiments.** Green highlight indicates identical V gene usage between the published sequence and the breastmilk-derived sequence found in this study. Blue highlight indicates concordance between previously published HLA epitope restriction and the participant's HLA type. For participants 3 and 6, no PBMC or BMC samples were available for HLA typing.

| Participant | Blood-derived CDR3b amino acid sequence | V gene of reference sequence | V gene of blood-derived TCR (this study) | Cognate Spike-specific epitope | Epitope HLA restriction (reference) | Relevant HLA allele of participant (this study) | Templates (this study) | Productive frequency (this study) | Reference PMID |
|---|---|---|---|---|---|---|---|---|---|
| 1 | CASGGTNTGELFF | TRBV2 | TCRBV12-03/12-04*01 | YLQPRTFLL | A*02:01 | None | 1 | 1.1757E-05 | 33326767 |
| 1 | CASSDLDTGELFF | TRBV2 | TCRBV06-04*01 | YLQPRTFLL | A*02:01 | None | 1 | 1.1757E-05 | 33326767 |
| | | TRBV2 | | YLQPRTFLL | A*02:01 | None | | | 34341799 |
| | | TRBV2 | | YLQPRTFLL | A*02:01 | None | | | 33951417 |
| 1 | CASSEANTGELFF | TRBV7-9 | TCRBV06-04*01 | YLQPRTFLL | A*02:01 | None | 1 | 1.1757E-05 | 33326767 |
| 1 | CASSETGGYEQYF | TRBV6-4 | TCRBV06-02*02 | NYNYLYRLF | A*24:02 | None | 1 | 1.1757E-05 | 34341799 |
| | | TRBV6-1 | | NYNYLYRLF | A*24:02 | | | | 34341799 |
| | | TRBV4-1 | | NYNYLYRLF | A*24:02 | | | | 34341799 |
| 1 | CASSGLNTGELFF | TRBV5-1 | TCRBV18-01*01 | YLQPRTFLL | A*02:01 | None | 1 | 1.1757E-05 | 33326767 |
| | | TRBV5-1 | | YLQPRTFLL | A*02:01 | | | | 33951417 |
| 1 | CASSLAGPNEQFF | TRBV7-9 | TCRBV07-07*01 | YLQPRTFLL | A*02:01 | None | 1 | 1.1757E-05 | 33951417 |
| 1 | CASSLAGYEQYF | TRBV5-1 | TCRBV28-01*01 | LTDEMIAQY | A*01:01 | A*01:01:01 | 1 | 1.1757E-05 | 34341799 |
| 1 | CASSLAQGYEQYF | TRBV12-3 | TCRBV07-09*01 | NYNYLYRLF | A*24:02 | None | 2 | 2.35139E-05 | 34341799 |
| 1 | CASSLASTDTQYF | TRBV28 | TCRBV28-01*01 | YLQPRTFLL | A*02:01 | None | 1 | 1.1757E-05 | 34341799 |
| 1 | CASSLAVNTEAFF | TRBV27 | TCRBV05-06*01 | LTDEMIAQY | A*01:01 | A*01:01:01 | 1 | 1.1757E-05 | 34341799 |
| 1 | CASSLGGNQPQHF | TRBV12-3 | TCRBV05-06*01 | LTDEMIAQY | A*01:01 | A*01:01:01 | 2 | 2.35139E-05 | 34341799 |
| 1 | CASSLGGNQPQHF | | TCRBV28-01*01 | | | | 1 | 1.1757E-05 | |
| 1 | CASSLGGNQPQHF | | TCRBV12-03/12-04*01 | | | | 2 | 2.35139E-05 | |
| 1 | CASSLGPEAFF | TRBV7-9 | TCRBV06-05*01 | YLQPRTFLL | A*02:01 | None | 1 | 1.1757E-05 | 34341799 |
| 1 | CASSLGTGYQPQHF | TRBV6-5 | TCRBV11-02*01 | RLQSLQTYV | A*02:01 | None | 1 | 1.1757E-05 | 33326767 |

| | | | | | | | | | |
|---|---|---|---|---|---|---|---|---|---|
| 1 | CASSLVQGGNTGELFF | TRBV13 | TCRBV11-02 | YLQPRTFLL | A*02:01 | None | 1 | 1.1757E-05 | 33326767 |
| 1 | CASSPGQGYEQYF | TRBV6-4 | TCRBV06-06*04 | NYNYLYRLF | A*24:02 | None | 1 | 1.1757E-05 | 34341799 |
| 1 | CASSPLSYEQYF | TRBV12-4 | TCRBV18-01*01 | LTDEMIAQY | A*01:01 | A*01:01:01 | 1 | 1.1757E-05 | 34341799 |
| 1 | CASSRVWDTEAFF | TRBV2 | TCRBV06-05*01 | AEVQIDRLI | B*44:02 | None | 1 | 1.1757E-05 | 34341799 |
| 1 | CASSVAGSSYEQYF | TRBV2 | TCRBV09-01*01 | LTDEMIAQY | A*01:01 | A*01:01:01 | 3 | 3.52709E-05 | 34341799 |
| 1 | CASSYGNQPQHF | TRBV7-9 | TCRBV06-06*01 | YLQPRTFLL | A*02:01 | None | 2 | 2.35139E-05 | 34341799 |
| 2 | CASGEANTGELFF | TRBV12-3 | TCRBV12-03/12-04*01 | YLQPRTFLL | A*02:01 | A*02:01:01 | 1 | 1.26229E-05 | 33326767 |
| 2 | CASGEENTGELFF | TRBV5-1 | TCRBV05-01*01 | YLQPRTFLL | A*02:01 | A*02:01:01 | 5 | 6.31146E-05 | 33326767 |
| 2 | CASLRDMNTGELFF | TRBV4-1 | TCRBV02-01 | YLQPRTFLL | A*02:01 | A*02:01:01 | 2 | 2.52458E-05 | 33326767 |
| 2 | CASQDLNTGELFF | TRBV19 | TCRBV02-01 | YLQPRTFLL | A*02:01 | A*02:01:01 | 2 | 2.52458E-05 | 33326767 |
| | | TRBV10-1 | | YLQPRTFLL | A*02:01 | A*02:01:01 | | | 33326767 |
| 2 | CASQDTNTGELFF | TRBV2 | TCRBV19-01*01 | YLQPRTFLL | A*02:01 | A*02:01:01 | 1 | 1.26229E-05 | 33951417 |
| 2 | CASQDTNTGELFF | | TCRBV07-03*01 | | | | 4 | 5.04917E-05 | |
| 2 | CASQEANTGELFF | TRBV19 | TCRBV06-05*01 | YLQPRTFLL | A*02:01 | A*02:01:01 | 2 | 2.52458E-05 | 33326767 |
| 2 | CASQGLNTGELFF | TRBV15 | TCRBV27-01*01 | YLQPRTFLL | A*02:01 | A*02:01:01 | 1 | 1.26229E-05 | 34341799 |
| | | TRBV15 | | YLQPRTFLL | A*02:01 | A*02:01:01 | | | 33326767 |
| 2 | CASSADIEAFF | TRBV7-9 | TCRBV07-09*01 | YLQPRTFLL | A*02:01 | A*02:01:01 | 1 | 1.26229E-05 | 34341799 |
| 2 | CASSDTNTGELFF | TRBV5-1 | TCRBV06-04*01 | YLQPRTFLL | A*02:01 | A*02:01:01 | 185 | 0.002335239 | 33326767 |
| 2 | CASSELNTGELFF | TRBV7-8 | TCRBV02-01 | YLQPRTFLL | A*02:01 | A*02:01:01 | 2 | 2.52458E-05 | 33326767 |
| | | TRBV2 | | YLQPRTFLL | A*02:01 | A*02:01:01 | | | 33664060 |
| 2 | CASSEQNTGELFF | TRBV9 | TCRBV10-02*01 | YLQPRTFLL | A*02:01 | A*02:01:01 | 1 | 1.26229E-05 | 34341799 |
| 2 | CASSFQNTGELFF | TRBV7-8 | TCRBV05-01*01 | YLQPRTFLL | A*02:01 | A*02:01:01 | 1 | 1.26229E-05 | 33326767 |
| 2 | CASSFTSSYNEQFF | TRBV28 | TCRBV05-01*01 | NQKLIANQF | B*15:01 | B*15:17:01 | 1 | 1.26229E-05 | 34341799 |
| 2 | CASSGTSGSTDTQYF | TRBV6-4 | TCRBV06-04*01 | NQKLIANQF | B*15:01 | B*15:17:01 | 2 | 2.52458E-05 | 34341799 |

| | | | | | | | | | |
|---|---|---|---|---|---|---|---|---|---|
| 2 | CASSLAGYEQYF | TRBV5-1 | TCRBV11-02*03 | LTDEMIAQY | A*01:01 | None | 1 | 1.26229E-05 | 34341799 |
| 2 | CASSLAGYEQYF | | TCRBV05-01*01 | | | | 1 | 1.26229E-05 | |
| 2 | CASSLAVNTEAFF | TRBV27 | TCRBV28-01*01 | LTDEMIAQY | A*01:01 | None | 1 | 1.26229E-05 | 34341799 |
| 2 | CASSLAVNTEAFF | | TCRBV05-01*01 | | | | 1 | 1.26229E-05 | |
| 2 | CASSLDIEAFF | TRBV7-9 | TCRBV07-06*01 | YLQPRTFLL | A*02:01 | A*02:01:01 | 1 | 1.26229E-05 | 34341799 |
| 2 | CASSLDIEAFF | TRBV7-9 | TCRBV07-09*01 | YLQPRTFLL | A*02:01 | A*02:01:01 | 14 | 0.000176721 | 33326767 |
| | | TRBV7-9 | | YLQPRTFLL | A*02:01 | A*02:01:01 | | | 33951417 |
| 2 | CASSLDIEQYF | TRBV7-9 | TCRBV07-09*01 | YLQPRTFLL | A*02:01 | A*02:01:01 | 1 | 1.26229E-05 | 34341799 |
| | | TRBV7-9 | | YLQPRTFLL | A*02:01 | A*02:01:01 | | | 33951417 |
| 2 | CASSLDVEQYF | TRBV7-9 | TCRBV07-09*01 | YLQPRTFLL | A*02:01 | A*02:01:01 | 1 | 1.26229E-05 | 33326767 |
| 2 | CASSLGGNQPQHF | TRBV12-3 | TCRBV05-01*01 | YLQPRTFLL | A*02:01 | A*02:01:01 | 1 | 1.26229E-05 | 34341799 |
| | CASSLGGNQPQHF | | TCRBV07-02 | | | | 1 | 1.26229E-05 | |
| 2 | CASSLGTGHQPQHF | TRBV27 | TCRBV27-01*01 | RLQSLQTYV | A*02:01 | A*02:01:01 | 2 | 2.52458E-05 | 33326767 |
| 2 | CASSLTGGGYTF | TRBV12-4 | TCRBV06-05*01 | NQKLIANQF | B*15:01 | B*15:17:01 | 1 | 1.26229E-05 | 34341799 |
| 2 | CASSLTSGAYNEQFF | TRBV19 | TCRBV18-01*01 | NQKLIANQF | B*15:01 | B*15:17:01 | 1 | 1.26229E-05 | 34341799 |
| 2 | CASSLVAGGNTGELFF | TRBV13 | TCRBV05-01*01 | YLQPRTFLL | A*02:01 | A*02:01:01 | 1 | 1.26229E-05 | 33326767 |
| 2 | CASSLYGGYEQYF | TRBV7-3 | TCRBV12-03/12-04*01 | YLQPRTFLL | A*02:01 | A*02:01:01 | 1 | 1.26229E-05 | 34341799 |
| 2 | CASSNNYEQYF | TRBV19 | TCRBV05-01*01 | NYNYLYRLF | A*24:02 | None | 1 | 1.26229E-05 | 34341799 |
| 2 | CASSPDIEAFF | TRBV7-9 | TCRBV07-09*01 | YLQPRTFLL | A*02:01 | A*02:01:01 | 6 | 7.57375E-05 | 33326767 |
| | | TRBV7-9 | | YLQPRTFLL | A*02:01 | | | | 34341799 |
| | | TRBV7-9 | | YLQPRTFLL | A*02:01 | | | | 33951417 |
| 2 | CASSPDIEQYF | TRBV7-9 | TCRBV07-09*01 | YLQPRTFLL | A*02:01 | A*02:01:01 | 3 | 3.78687E-05 | 33326767 |
| | | TRBV7-9 | | YLQPRTFLL | A*02:01 | | | | 34341799 |
| | | TRBV7-9 | | YLQPRTFLL | A*02:01 | | | | 33951417 |
| 2 | CASSPEIEAFF | TRBV7-9 | TCRBV07-09*01 | YLQPRTFLL | A*02:01 | A*02:01:01 | 35 | 0.000441802 | 33326767 |

| | | | | | | | | | |
|---|---|---|---|---|---|---|---|---|---|
| 2 | CASSPGDTQYF | TRBV25-1 | TCRBV12-03/12-04*01 | YLQPRTFLL | A*02:01 | A*02:01:01 | 1 | 1.26229E-05 | 33951417 |
| 2 | CASSPGDTQYF | | TCRBV07-03*01 | | | | 1 | 1.26229E-05 | |
| 2 | CASSPGQGYEQYF | TRBV6-4 | TCRBV13-01*01 | NYNYLYRLF | A*24:02 | None | 2 | 2.52458E-05 | 34341799 |
| 2 | CASSPPGGGNTGELFF | TRBV18 | TCRBV05-08*01 | YLQPRTFLL | A*02:01 | A*02:01:01 | 30 | 0.000378687 | 33326767 |
| 2 | CASSQDLQGTNEKLFF | TRBV4-2 | TCRBV04-02*01 | RLQSLQTYV | A*02:01 | A*02:01:01 | 3 | 3.78687E-05 | 33326767 |
| 2 | CASSRLAGGRNEQFF | TRBV5-4 | TCRBV12 | YLQPRTFLL | A*02:01 | A*02:01:01 | 1 | 1.26229E-05 | 33951417 |
| 2 | CASSSDIEAFF | TRBV7-9 | TCRBV07-09*01 | YLQPRTFLL | A*02:01 | A*02:01:01 | 1 | 1.26229E-05 | 33326767 |
| 2 | CAWSVQQNYGYTF | | TCRBV30-01*01 | SKRSFIEDL<br>LFNKVTLA | DPA1*01:03/<br>DPB1*04:01 | Unknown | 1 | 1.26229E-05 | 33830946 |
| 2 | CSARDRQGQNTGELFF | TRBV20-1 | TCRBV20 | YLQPRTFLL | A*02:01 | A*02:01:01 | 1 | 1.26229E-05 | 33951417 |
| 4 | CASSDANTGELFF | TRBV5-1 | TCRBV06-04*01 | YLQPRTFLL | A*02:01 | None | 3 | 5.61545E-05 | 33326767 |
| | | TRBV7-9 | | YLQPRTFLL | A*02:01 | None | | | 34341799 |
| 4 | CASSDGNTGELFF | TRBV5-1 | TCRBV06-04*01 | YLQPRTFLL | A*02:01 | None | 2 | 3.74364E-05 | 33326767 |
| 4 | CASSDLNTGELFF | TRBV2 | TCRBV06-04*01 | YLQPRTFLL | A*02:01 | None | 1 | 1.87182E-05 | 33326767 |
| | | TRBV2 | | YLQPRTFLL | A*02:01 | None | | | 33951417 |
| 4 | CASSDTNTGELFF | TRBV5-1 | TCRBV06-04*01 | YLQPRTFLL | A*02:01 | None | 1 | 1.87182E-05 | 33326767 |
| 4 | CASSLAQGYEQYF | TRBV12-3 | TCRBV11-03*01 | NYNYLYRLF | A*24:02 | A*24:02:01 | 1 | 1.87182E-05 | 34341799 |
| 4 | CASSLASNQPQHF | TRBV27 | TCRBV07-06*01 | LTDEMIAQY | A*01:01 | None | 1 | 1.87182E-05 | 34341799 |
| 4 | CASSLGGNQPQHF | TRBV12-3 | TCRBV07-08*01 | LTDEMIAQY | A*01:01 | None | 1 | 1.87182E-05 | 34341799 |
| 4 | CASSLGGNQPQHF | TRBV12-3 | TCRBV12-03/12-04*01 | LTDEMIAQY | A*01:01 | None | 1 | 1.87182E-05 | 34341799 |
| 4 | CASSPGQGYEQYF | TRBV6-4 | TCRBV12-03/12-04*01 | NYNYLYRLF | A*24:02 | A*24:02:01 | 2 | 3.74364E-05 | 34341799 |

| | | | | | | | | | |
|---|---|---|---|---|---|---|---|---|---|
| 4 | CASSQGQGNIQYF | TRBV4-3 | TCRBV04-03*01 | RLQSLQTYV | A*02:01 | None | 1 | 1.87182E-05 | 33326767 |
| 4 | CASSQGSGNEQFF | TRBV5-6 | TCRBV04-01*01 | QYIKWPWYI | A*24:02 | A*24:02:01 | 1 | 1.87182E-05 | 34341799 |
| 4 | CAWSVQGNYGYTF | | TCRBV30-01*01 | SKRSFIEDL LFNKVTLA | DPA1*01:03/ DPB1*04:01 | Unknown | 1 | 1.87182E-05 | 33830946 |
| 5 | CASQDSNTGELFF | TRBV12-3 | TCRBV10-01*01 | YLQPRTFLL | A*02:01 | None | 1 | 1.00112E-05 | 33326767 |
| 5 | CASRRGFEQYF | TRBV2 | TCRBV28-01*01 | YLQPRTFLL | A*02:01 | None | 1 | 1.00112E-05 | 33326767 |
| 5 | CASRSGQRNTEAFF | TRBV6-5 | TCRBV19-01*01 | LTDEMIAQY | A*01:01 | None | 1 | 1.00112E-05 | 34341799 |
| 5 | CASRSGTGTYEQYF | TRBV19 | TCRBV13-01*01 | LTDEMIAQY | A*01:01 | None | 1 | 1.00112E-05 | 34341799 |
| 5 | CASSDSYGYTF | TRBV7-8 | TCRBV25-01*01 | YLQPRTFLL | A*02:01 | None | 1 | 1.00112E-05 | 33951417 |
| | | | | | | None | | | 33326767 |
| 5 | CASSLAGYEQYF | TRBV5-1 | TCRBV05-01*01 | LTDEMIAQY | A*01:01 | None | 1 | 1.00112E-05 | 34341799 |
| 5 | CASSLASNQPQHF | TRBV6-5 | TCRBV11-01*01 | LTDEMIAQY | A*01:01 | None | 1 | 1.00112E-05 | 34341799 |
| 5 | CASSLAVNTEAFF | TRBV5-1 | TCRBV07-02 | LTDEMIAQY | A*01:01 | None | 1 | 1.00112E-05 | 34341799 |
| 5 | CASSLGDSYNEQFF | TRBV7-9 | TCRBV05-01*01 | LTDEMIAQY | A*01:01 | None | 2 | 2.00224E-05 | 34341799 |
| 5 | CASSLGGNQPQHF | TRBV12-3 | TCRBV07 | LTDEMIAQY | A*01:01 | None | 1 | 1.00112E-05 | 34341799 |
| 5 | CASSLGGNQPQHF | | TCRBV07-09*01 | | | None | 2 | 2.00224E-05 | |
| 5 | CASSLGGNQPQHF | | TCRBV12-03/12-04*01 | | | None | 1 | 1.00112E-05 | |
| 5 | CASSLGGNQPQHF | | TCRBV11-02*01 | | | None | 1 | 1.00112E-05 | |
| 5 | CASSLGPEAFF | TRBV7-9 | TCRBV05-06*01 | YLQPRTFLL | A*02:01 | None | 1 | 1.00112E-05 | 34341799 |
| 5 | CASSLGVGRAYEQYF | TRBV7-2 | TCRBV11-02*01 | NQKLIANQF | B*15:01 | None | 1 | 1.00112E-05 | 34341799 |
| 5 | CASSLQNTGELFF | TRBV7-8 | TCRBV05-06*01 | YLQPRTFLL | A*02:01 | None | 4 | 4.00449E-05 | 33951417 |
| 5 | CASSLTGGGYTF | TRBV12-4 | TCRBV12-03/12-04*01 | NQKLIANQF | B*15:01 | None | 1 | 1.00112E-05 | 34341799 |
| 5 | CASSPGQGYEQYF | TRBV6-4 | TCRBV21-01*01 | NYNYLYRLF | A*24:02 | None | 1 | 1.00112E-05 | 34341799 |

| | | | | | | | | | |
|---|---|---|---|---|---|---|---|---|---|
| 5 | CASSPGTSGGSSYNEQFF | TRBV11-2 | TCRBV07-03*01 | NQKLIANQF | B*15:01 | None | 1 | 1.00112E-05 | 34341799 |
| 5 | CASSPLSYEQYF | TRBV12-4 | TCRBV07-02 | LTDEMIAQY | A*01:01 | None | 1 | 1.00112E-05 | 34341799 |
| 5 | CASSQGGNEQYF | TRBV4-1 | TCRBV07 | YLQPRTFLL | A*02:01 | None | 1 | 1.00112E-05 | 34341799 |
| 5 | CAWSVQQNYGYTF | | TCRBV30-01*01 | SKRSFIEDL LFNKVTLA | DPA1*01:03/ DPB1*04:01 | Unknown | 1 | 1.00112E-05 | 33830946 |
| 7 | CASREGNTGELFF | TRBV4-2 | TCRBV09-01*01 | YLQPRTFLL | A*02:01 | None | 1 | 1.12293E-05 | 33326767 |
| 7 | CASREGNTGELFF | | TCRBV06-01*01 | | | None | 1 | 1.12293E-05 | |
| 7 | CASSDSYGYTF | TRBV7-8 | TCRBV09-01*01 | YLQPRTFLL | A*02:01 | None | 1 | 1.12293E-05 | 33326767 |
| | | | | | | None | | | 33951417 |
| 7 | CASSEAANQPQHF | TRBV27 | TCRBV06-01*01 | LTDEMIAQY | A*01:01 | None | 1 | 1.12293E-05 | 34341799 |
| 7 | CASSEGQGYEQYF | TRBV2 | TCRBV04-01*01 | NYNYLYRLF | A*24:02 | None | 1 | 1.12293E-05 | 34341799 |
| 7 | CASSGGNTIYF | TRBV28 | TCRBV06-05*01 | LTDEMIAQY | A*01:01 | None | 1 | 1.12293E-05 | 34341799 |
| 7 | CASSGGNTIYF | | TCRBV06 | | | None | 3 | 3.36878E-05 | |
| 7 | CASSHTNTGELFF | TRBV9 | TCRBV07 | YLQPRTFLL | A*02:01 | None | 1 | 1.12293E-05 | 33326767 |
| 7 | CASSKGGSNQPQHF | TRBV19 | TCRBV07-06*01 | NQKLIANQF | B*15:01 | None | 1 | 1.12293E-05 | 34341799 |
| 7 | CASSLAQGYEQYF | TRBV12-3 | TCRBV07-08*01 | NYNYLYRLF | A*24:02 | None | 1 | 1.12293E-05 | 34341799 |
| 7 | CASSLASTDTQYF | TRBV28 | TCRBV07-06*01 | YLQPRTFLL | A*02:01 | None | 1 | 1.12293E-05 | 34341799 |
| 7 | CASSLGGNQPQHF | TRBV12-3 | TCRBV12-03/12-04*01 | LTDEMIAQY | A*01:01 | None | 1 | 1.12293E-05 | 34341799 |
| 7 | CASSLGGNQPQHF | | TCRBV27-01*01 | | | None | 1 | 1.12293E-05 | |
| 7 | CASSLGGNQPQHF | | TCRBV06-05*01 | | | None | 1 | 1.12293E-05 | |
| 7 | CASSLQNTGELFF | TRBV7-8 | TCRBV05-01*01 | YLQPRTFLL | A*02:01 | None | 1 | 1.12293E-05 | 33951417 |
| 7 | CASSLTSGAYNEQFF | TRBV19 | TCRBV11-02*01 | NQKLIANQF | B*15:01 | None | 1 | 1.12293E-05 | 34341799 |
| 7 | CASSLVVTGELFF | TRBV7-3 | TCRBV05-01*01 | LTDEMIAQY | A*01:01 | None | 2 | 2.24585E-05 | 34341799 |

| | | | | | | | | | |
|---|---|---|---|---|---|---|---|---|---|
| 7 | CASSPDRNTGELFF | TRBV5-4 | TCRBV18-01*01 | YLQPRTFLL | A*02:01 | None | 2 | 2.24585E-05 | 33951417 |
| 7 | CASSPDRNTGELFF | | TCRBV02-01 | | | None | 1 | 1.12293E-05 | |
| 7 | CASSPGQGYEQYF | TRBV6-4 | TCRBV06-05*01 | NYNYLYRLF | A*24:02 | None | 1 | 1.12293E-05 | 34341799 |
| 7 | CASSPGQGYEQYF | | TCRBV07-03*01 | | | None | 1 | 1.12293E-05 | |
| 7 | CASSQGPNGYTF | TRBV4-1 | TCRBV03-01/03-02*01 | YLQPRTFLL | A*02:01 | None | 1 | 1.12293E-05 | 34341799 |
| 7 | CASSYGNQPQHF | TRBV7-9 | TCRBV06-02/06-03*01 | YLQPRTFLL | A*02:01 | None | 1 | 1.12293E-05 | 34341799 |
| 7 | CASSYQNTGELFF | TRBV7-8 | TCRBV05-01*01 | YLQPRTFLL | A*02:01 | None | 1 | 1.12293E-05 | 33326767 |
| 8 | CASSLAQGYEQYF | TRBV12-3 | TCRBV07-07*01 | NYNYLYRLF | A*24:02 | None | 1 | 1.44101E-05 | 34341799 |
| 8 | CASSLVVTGELFF | TRBV7-3 | TCRBV05-06*01 | LTDEMIAQY | A*01:01 | A*01:01:01 | 1 | 1.44101E-05 | 34341799 |
| 8 | CASSPLAGGNTGELFF | TRBV3-1 | TCRBV07-09*01 | YLQPRTFLL | A*02:01 | A*02:01:01 | 1 | 1.44101E-05 | 33326767 |
| 8 | CASSLASTDTQYF | TRBV28 | TCRBV05-01*01 | YLQPRTFLL | A*02:01 | A*02:01:01 | 1 | 1.44101E-05 | 34341799 |
| 8 | CASSLDSEQFF | TRBV7-9 | TCRBV05-01*01 | YLQPRTFLL | A*02:01 | A*02:01:01 | 1 | 1.44101E-05 | 33326767 |
| 8 | CASSYQNTGELFF | TRBV7-8 | TCRBV10-02*01 | YLQPRTFLL | A*02:01 | A*02:01:01 | 1 | 1.44101E-05 | 33326767 |
| 8 | CASSEQNTGELFF | TRBV9 | TCRBV10-02*01 | YLQPRTFLL | A*02:01 | A*02:01:01 | 1 | 1.44101E-05 | 34341799 |
| 8 | CASSDGNTGELFF | TRBV5-1 | TCRBV06-04*01 | YLQPRTFLL | A*02:01 | A*02:01:01 | 1 | 1.44101E-05 | 33326767 |
| 8 | CASSDTNTGELFF | TRBV5-1 | TCRBV06-04*01 | YLQPRTFLL | A*02:01 | A*02:01:01 | 18 | 0.000259381 | 33326767 |
| 8 | CASSEANTGELFF | TRBV7-9 | TCRBV06-04*01 | YLQPRTFLL | A*02:01 | A*02:01:01 | 2 | 2.88201E-05 | 33326767 |
| 8 | CASSYWGGTDTQYF | TRBV6-6 | TCRBV06-05*01 | LTDEMIAQY | A*01:01 | A*01:01:01 | 1 | 1.44101E-05 | 34341799 |
| 8 | CASSDSYGYTF | TRBV7-8 | TCRBV06-02/06-03*01 | YLQPRTFLL | A*02:01 | A*02:01:01 | 1 | 1.44101E-05 | 33951417 |
| | | TRBV7-8 | | YLQPRTFLL | A*02:01 | A*02:01:01 | | | 33326767 |
| 8 | CASSVAGSSYEQYF | TRBV2 | TCRBV09-01*01 | LTDEMIAQY | A*01:01 | A*01:01:01 | 1 | 1.44101E-05 | 34341799 |
| 8 | CASGTANTGELFF | TRBV12-3 | TCRBV10-01*01 | YLQPRTFLL | A*02:01 | A*02:01:01 | 1 | 1.44101E-05 | 33326767 |
| 8 | CASSDLNTGELFF | TRBV2 | TCRBV02-01 | YLQPRTFLL | A*02:01 | A*02:01:01 | 1 | 1.44101E-05 | 33951417 |

| | | | | | | | | | |
|---|---|---|---|---|---|---|---|---|---|
| | | TRBV2 | | YLQPRTFLL | A*02:01 | A*02:01:01 | | | 33326767 |
| 8 | CASSLAVNTEAFF | TRBV27 | TCRBV07-02 | LTDEMIAQY | A*01:01 | A*01:01:01 | 1 | 1.44101E-05 | 34341799 |
| 8 | CASSPGSYSNQPQHF | TRBV9 | TCRBV02-01 | NQKLIANQF | B*15:01 | None | 1 | 1.44101E-05 | 34341799 |
| 8 | CASSELNTGELFF | TRBV2 | TCRBV02-01 | YLQPRTFLL | A*02:01 | A*02:01:01 | 3 | 4.32302E-05 | 33664060 |
| | | TRBV7-8 | | YLQPRTFLL | A*02:01 | A*02:01:01 | | | |
| 9 | CASSDGNTGELFF | TRBV5-1 | TCRBV06-04*01 | YLQPRTFLL | A*02:01 | None | 1 | 1.42542E-05 | 33326767 |
| 9 | CASSDGNTGELFF | | TCRBV11-03*01 | | | None | 1 | 1.42542E-05 | |
| 9 | CASSDSYGYTF | TRBV7-8 | TCRBV06-05*01 | YLQPRTFLL | A*02:01 | None | 1 | 1.42542E-05 | 33326767 |
| | | TRBV7-8 | | YLQPRTFLL | A*02:01 | None | | | 33951417 |
| 9 | CASSDTNTGELFF | TRBV5-1 | TCRBV06-04*01 | YLQPRTFLL | A*02:01 | None | 5 | 7.12708E-05 | 33326767 |
| 9 | CASSEAQGYEQYF | TRBV7-2 | TCRBV02-01 | NYNYLYRLF | A*24:02 | None | 1 | 1.42542E-05 | 34341799 |
| 9 | CASSEGRGYEQYF | TRBV6-1 | TCRBV06-04*01 | NYNYLYRLF | A*24:02 | None | 1 | 1.42542E-05 | 34341799 |
| 9 | CASSENRGYEQYF | TRBV2 | TCRBV06 | NYNYLYRLF | A*24:02 | None | 2 | 2.85083E-05 | 34341799 |
| 9 | CASSFQNTGELFF | TRBV7-8 | TCRBV07-02 | YLQPRTFLL | A*02:01 | None | 1 | 1.42542E-05 | 33326767 |
| 9 | CASSGTSGSTDTQYF | TRBV6-4 | TCRBV06-04*01 | NQKLIANQF | B*15:01 | None | 1 | 1.42542E-05 | 34341799 |
| 9 | CASSLAGYEQYF | TRBV5-1 | TCRBV07-09*01 | LTDEMIAQY | A*01:01 | A*01:01:01 | 1 | 1.42542E-05 | 34341799 |
| 9 | CASSLAGYEQYF | | TCRBV11-02*01 | | | A*01:01:01 | 1 | 1.42542E-05 | |
| 9 | CASSLASNQPQHF | TRBV27 | TCRBV05-06*01 | LTDEMIAQY | A*01:01 | A*01:01:01 | 1 | 1.42542E-05 | 34341799 |
| 9 | CASSLASNQPQHF | | TCRBV07 | | | A*01:01:01 | 1 | 1.42542E-05 | |
| 9 | CASSLAVNTEAFF | TRBV27 | TCRBV11-03*01 | LTDEMIAQY | A*01:01 | A*01:01:01 | 1 | 1.42542E-05 | 34341799 |
| 9 | CASSLEDTYYGYTF | TRBV7-2 | TCRBV05-06*01 | NQKLIANQF | B*15:01 | None | 1 | 1.42542E-05 | 34341799 |
| 9 | CASSLGDSYNEQFF | TRBV7-9 | TCRBV05-01*01 | LTDEMIAQY | A*01:01 | A*01:01:01 | 1 | 1.42542E-05 | 34341799 |
| 9 | CASSLGGNQPQHF | TRBV12-3 | TCRBV05-06*01 | LTDEMIAQY | A*01:01 | A*01:01:01 | 3 | 4.27625E-05 | 34341799 |
| 9 | CASSLGGNQPQHF | | TCRBV28-01*01 | | | A*01:01:01 | 1 | 1.42542E-05 | |
| 9 | CASSLGGNQPQHF | | TCRBV05-01*01 | | | A*01:01:01 | 1 | 1.42542E-05 | |
| 9 | CASSNNYEQYF | TRBV19 | TCRBV21-01*01 | NYNYLYRLF | A*24:02 | None | 1 | 1.42542E-05 | 34341799 |
| 9 | CASSPDRNTGELFF | TRBV5-4 | TCRBV05-01*01 | YLQPRTFLL | A*02:01 | None | 2 | 2.85083E-05 | 33951417 |

| | | | | | | | | | |
|---|---|---|---|---|---|---|---|---|---|
| 9 | CASSPDSEQYF | TRBV7-9 | TCRBV02-01 | YLQPRTFLL | A*02:01 | None | 1 | 1.42542E-05 | 33326767 |
| 9 | CASSPGQGYEQYF | TRBV6-4 | TCRBV03-01/03-02*01 | NYNYLYRLF | A*24:02 | None | 1 | 1.42542E-05 | 33326767 |
| 9 | CASSPGQGYEQYF | | TCRBV18-01*01 | | | None | 1 | 1.42542E-05 | 33326767 |
| 9 | CASSPGQGYEQYF | | TCRBV28-01*01 | | | None | 1 | 1.42542E-05 | 33326767 |
| 9 | CASSPGQGYEQYF | | TCRBV12-03/12-04*01 | | | None | 1 | 1.42542E-05 | 33326767 |
| 9 | CASSPLSYEQYF | TRBV12-4 | TCRBV18-01*01 | LTDEMIAQY | A*01:01 | A*01:01:01 | 1 | 1.42542E-05 | 34341799 |
| 9 | CASSQGSGNEQFF | TRBV5-6 | TCRBV04-03*01 | QYIKWPWYI | A*24:02 | None | 1 | 1.42542E-05 | 34341799 |
| 9 | CASSRDIEAFF | TRBV7-9 | TCRBV07-09*01 | YLQPRTFLL | A*02:01 | None | 1 | 1.42542E-05 | 34341799 |
| 9 | CASSSLNTGELFF | TRBV2 | TCRBV07-09*01 | YLQPRTFLL | A*02:01 | None | 1 | 1.42542E-05 | 33326767 |
| 9 | CATGLANTGELFF | TRBV7-9 | TCRBV24-01*01 | YLQPRTFLL | A*02:01 | None | 17 | 0.000242321 | 34341799 |
| 10 | CASREGNTGELFF | TRBV4-2 | TCRBV19-01*01 | YLQPRTFLL | A*02:01 | None | 1 | 9.14854E-06 | 33326767 |
| | CASREGNTGELFF | | TCRBV28-01*01 | | | None | 1 | 9.14854E-06 | |
| 10 | CASSFLAGGNTGELFF | TRBV19 | TCRBV03-01/03-02*01 | YLQPRTFLL | A*02:01 | None | 1 | 9.14854E-06 | 33326767 |
| 10 | CASSGGNTIYF | TRBV28 | TCRBV28-01*01 | LTDEMIAQY | A*01:01 | None | 1 | 9.14854E-06 | 34341799 |
| 10 | CASSGGNTIYF | | TCRBV06-05*01 | | | None | 1 | 9.14854E-06 | |
| 10 | CASSGTSGSTDTQYF | TRBV6-4 | TCRBV06-04*01 | NQKLIANQF | B*15:01 | None | 1 | 9.14854E-06 | 34341799 |
| 10 | CASSLAGYEQYF | TRBV5-1 | TCRBV05-06*01 | LTDEMIAQY | A*01:01 | None | 2 | 1.82971E-05 | 34341799 |
| 10 | CASSLAQGYEQYF | TRBV12-3 | TCRBV18-01*01 | NYNYLYRLF | A*24:02 | None | 1 | 9.14854E-06 | 34341799 |
| 10 | CASSLGASSYNEQFF | TRBV12-3 | TCRBV07-03*01 | YLQPRTFLL | A*02:01 | None | 1 | 9.14854E-06 | 34341799 |
| 10 | CASSLGASSYNEQFF | | TCRBV07-02 | | | None | 1 | 9.14854E-06 | |
| 10 | CASSLGDSYNEQFF | TRBV7-9 | TCRBV05-01*01 | LTDEMIAQY | A*01:01 | None | 1 | 9.14854E-06 | 34341799 |
| 10 | CASSLGGNQPQHF | | TCRBV05-01*01 | LTDEMIAQY | A*01:01 | None | 1 | 9.14854E-06 | 34341799 |

| | | | | | | | | | |
|---|---|---|---|---|---|---|---|---|---|
| | CASSLGGNQPQHF | TRBV12-3 | TCRBV27-01*01 | | | None | 1 | 9.14854E-06 | |
| | CASSLGGNQPQHF | | TCRBV28-01*01 | | | None | 2 | 1.82971E-05 | |
| 10 | CASSLQNTGELFF | TRBV7-8 | TCRBV14-01*01 | YLQPRTFLL | A*02:01 | None | 2 | 1.82971E-05 | 33951417 |
| 10 | CASSLTSGAYNEQFF | TRBV19 | TCRBV06-05*01 | NQKLIANQF | B*15:01 | None | 1 | 9.14854E-06 | 34341799 |
| 10 | CASSPDSEQYF | TRBV7-9 | TCRBV07-09*01 | YLQPRTFLL | A*02:01 | None | 1 | 9.14854E-06 | 33326767 |
| 10 | CASSPGQGYEQYF | TRBV6-4 | TCRBV04-03*01 | NYNYLYRLF | A*24:02 | None | 1 | 9.14854E-06 | 34341799 |
| 10 | CASTRDIEAFF | TRBV7-9 | TCRBV25-01*01 | YLQPRTFLL | A*02:01 | None | 1 | 9.14854E-06 | 33326767 |
| 11 | CASEDRNTGELFF | TRBV6-6 | TCRBV06-06 | YLQPRTFLL | A*02:01 | None | 1 | 8.47443E-06 | 33326767 |
| 11 | CASRRDRAYEQYF | TRBV12-3 | TCRBV12-03/12-04*01 | NQKLIANQF | B*15:01 | None | 1 | 8.47443E-06 | 34341799 |
| 11 | CASSDGNTGELFF | TRBV5-1 | TCRBV02-01 | YLQPRTFLL | A*02:01 | None | 1 | 8.47443E-06 | 33326767 |
| 11 | CASSDGNTGELFF | | TCRBV10-01*01 | | | None | 1 | 8.47443E-06 | |
| 11 | CASSDLDTGELFF | TRBV2 | TCRBV10-01*01 | YLQPRTFLL | A*02:01 | None | 1 | 8.47443E-06 | 34341799 |
| 11 | CASSEANTGELFF | TRBV7-9 | TCRBV10-02*01 | YLQPRTFLL | A*02:01 | None | 1 | 8.47443E-06 | 33326767 |
| 11 | CASSEGQGYEQYF | TRBV5-1 | TCRBV02-01 | NYNYLYRLF | A*24:02 | None | 1 | 8.47443E-06 | 34341799 |
| 11 | CASSEGQGYEQYF | TRBV2 | TCRBV11-02*01 | NYNYLYRLF | A*24:02 | None | 1 | 8.47443E-06 | 34341799 |
| | | TRBV6-1 | | NYNYLYRLF | A*24:02 | None | | | 34341799 |
| 11 | CASSEQNTGELFF | TRBV9 | TCRBV07-03*01 | YLQPRTFLL | A*02:01 | None | 1 | 8.47443E-06 | 34341799 |
| 11 | CASSFTSSYNEQFF | TRBV28 | TCRBV06-02/06-03*01 | NQKLIANQF | B*15:01 | None | 1 | 8.47443E-06 | 34341799 |
| 11 | CASSFWGGGTEAFF | TRBV5-6 | TCRBV28-01*01 | NQKLIANQF | B*15:01 | None | 1 | 8.47443E-06 | 34341799 |
| 11 | CASSGGQGANTGELFF | TRBV2 | TCRBV05-06*01 | YLQPRTFLL | A*02:01 | None | 1 | 8.47443E-06 | 33951417 |
| 11 | CASSGTSGSTDTQYF | TRBV6-4 | TCRBV06-04*01 | NQKLIANQF | B*15:01 | None | 13 | 0.000110168 | 34341799 |
| 11 | CASSIGDEQYF | TRBV19 | TCRBV19-01*01 | YLQPRTFLL | A*02:01 | None | 1 | 8.47443E-06 | 33951417 |
| 11 | CASSLAGYEQYF | TRBV5-1 | TCRBV05-06*01 | LTDEMIAQY | A*01:01 | None | 2 | 1.69489E-05 | 34341799 |
| | CASSLAGYEQYF | | TCRBV28-01*01 | | | None | 1 | 8.47443E-06 | |
| | CASSLAGYEQYF | | TCRBV11-02*01 | | | None | 1 | 8.47443E-06 | |

| | | | | | | | | | |
|---|---|---|---|---|---|---|---|---|---|
| | CASSLAGYEQYF | | TCRBV07 | | | None | 1 | 8.47443E-06 | |
| 11 | CASSLAQGYEQYF | TRBV12-3 | TCRBV28-01*01 | NYNYLYRLF | A*24:02 | None | 1 | 8.47443E-06 | 34341799 |
| 11 | CASSLASTDTQYF | TRBV28 | TCRBV05-01*01 | YLQPRTFLL | A*02:01 | None | 1 | 8.47443E-06 | 34341799 |
| 11 | CASSLAVNTEAFF | TRBV27 | TCRBV05-06*01 | LTDEMIAQY | A*01:01 | None | 1 | 8.47443E-06 | 34341799 |
| 11 | CASSLDSEQFF | TRBV7-9 | TCRBV18-01*01 | YLQPRTFLL | A*02:01 | None | 1 | 8.47443E-06 | 33326767 |
| 11 | CASSLGAGTYEQYF | TRBV7-8 | TCRBV07-02 | LTDEMIAQY | A*01:01 | None | 1 | 8.47443E-06 | 34341799 |
| 11 | CASSLGDSYNEQFF | TRBV7-9 | TCRBV13-01*01 | LTDEMIAQY | A*01:01 | None | 1 | 8.47443E-06 | 34341799 |
| 11 | CASSLGGNQPQHF | TRBV12-3 | TCRBV07-02 | LTDEMIAQY | A*01:01 | None | 1 | 8.47443E-06 | 34341799 |
| | CASSLGGNQPQHF | | TCRBV11-02*01 | | | None | 1 | 8.47443E-06 | |
| | CASSLGGNQPQHF | | TCRBV28-01*01 | | | None | 1 | 8.47443E-06 | |
| | CASSLGGNQPQHF | | TCRBV07-09*01 | | | None | 1 | 8.47443E-06 | |
| 11 | CASSLGIAKNIQYF | TRBV11-2 | TCRBV11-02*01 | YLQPRTFLL | A*02:01 | None | 1 | 8.47443E-06 | 34341799 |
| 11 | CASSLGPEAFF | TRBV7-9 | TCRBV05-05*01 | YLQPRTFLL | A*02:01 | None | 1 | 8.47443E-06 | 34341799 |
| 11 | CASSLGTKNIQYF | TRBV4-2 | TCRBV05-04*01 | LTDEMIAQY | A*01:01 | None | 1 | 8.47443E-06 | 34341799 |
| 11 | CASSLIGGSSGNTIYF | TRBV2 | TCRBV28-01*01 | QYIKWPWYI | A*24:02 | None | 1 | 8.47443E-06 | 34341799 |
| 11 | CASSLQNTGELFF | TRBV7-8 | TCRBV11-03*01 | YLQPRTFLL | A*02:01 | None | 1 | 8.47443E-06 | 33951417 |
| 11 | CASSLTSGAYNEQFF | TRBV19 | TCRBV07-02 | NQKLIANQF | B*15:01 | None | 1 | 8.47443E-06 | 34341799 |
| 11 | CASSPDRGGGYTF | TRBV18 | TCRBV05-04*01 | YLQPRTFLL | A*02:01 | None | 1 | 8.47443E-06 | 34341799 |
| 11 | CASSPGQGYEQYF | TRBV6-4 | TCRBV19-01*01 | NYNYLYRLF | A*24:02 | None | 2 | 1.69489E-05 | 34341799 |
| 11 | CASSPPGGGNTGELFF | TRBV18 | TCRBV07-02 | YLQPRTFLL | A*02:01 | None | 1 | 8.47443E-06 | 33326767 |
| 11 | CASSQGGSADTQYF | TRBV3-1 | TCRBV05-04*01 | NQKLIANQF | B*15:01 | None | 1 | 8.47443E-06 | 34341799 |
| 11 | CASSSQNTGELFF | TRBV7-8 | TCRBV28-01*01 | YLQPRTFLL | A*02:01 | None | 1 | 8.47443E-06 | 33326767 |
| 11 | CASSTGGVGYEQYF | TRBV27 | TCRBV19-01*01 | NQKLIANQF | B*15:01 | None | 1 | 8.47443E-06 | 34341799 |
| 11 | CASSTRDSNQPQHF | TRBV19 | TCRBV27-01*01 | YLQPRTFLL | A*02:01 | None | 1 | 8.47443E-06 | 34341799 |
| 11 | CASSVAGSSYEQYF | TRBV2 | TCRBV09-01*01 | LTDEMIAQY | A*01:01 | None | 1 | 8.47443E-06 | 34341799 |

| | | | | | | | | | |
|---|---|---|---|---|---|---|---|---|---|
| 11 | CATSDSNTGELFF | TRBV5-1 | TCRBV24-01*01 | YLQPRTFLL | A*02:01 | None | 1 | 8.47443E-06 | 33951417 |
| 11 | CSARDRSSYEQYF | TRBV20-1 | TCRBV20 | YLQPRTFLL | A*02:01 | None | 1 | 8.47443E-06 | 34341799 |
| 12 | CASSDSYGYTF | TRBV7-8 | TCRBV06-05*01 | YLQPRTFLL | A*02:01 | None | 1 | 9.09314E-06 | 33326767 |
| | | TRBV7-8 | | YLQPRTFLL | A*02:01 | None | | | 33951417 |
| 12 | CASSEGQGYEQYF | TRBV5-1 | TCRBV05-01*01 | NYNYLYRLF | A*24:02 | None | 1 | 9.09314E-06 | 34341799 |
| 12 | CASSEGQGYEQYF | TRBV2 | TCRBV02-01 | NYNYLYRLF | A*24:02 | None | 1 | 9.09314E-06 | 34341799 |
| | | TRBV6-1 | | NYNYLYRLF | A*24:02 | None | | | 34341799 |
| 12 | CASSEGRGYEQYF | TRBV6-1 | TCRBV02-01 | NYNYLYRLF | A*24:02 | None | 1 | 9.09314E-06 | 34341799 |
| 12 | CASSEIDTGELFF | TRBV2 | TCRBV10-02*01 | YLQPRTFLL | A*02:01 | None | 1 | 9.09314E-06 | 33951417 |
| 12 | CASSEWIQETQYF | TRBV6-1 | TCRBV10-02*01 | YLQPRTFLL | A*02:01 | None | 1 | 9.09314E-06 | 33326767 |
| 12 | CASSGGQGANTGELFF | TRBV2 | TCRBV05-06*01 | YLQPRTFLL | A*02:01 | None | 19 | 0.00017277 | 33951417 |
| 12 | CASSGTSGSTDTQYF | TRBV6-4 | TCRBV19-01*01 | NQKLIANQF | B*15:01 | None | 1 | 9.09314E-06 | 34341799 |
| 12 | CASSLASTDTQYF | TRBV28 | TCRBV07-07*01 | YLQPRTFLL | A*02:01 | None | 1 | 9.09314E-06 | 34341799 |
| 12 | CASSLAVNTEAFF | TRBV27 | TCRBV05-01*01 | LTDEMIAQY | A*01:01 | A*01:01:01 | 1 | 9.09314E-06 | 34341799 |
| 12 | CASSLGASSYNEQFF | TRBV12-3 | TCRBV05-01*01 | YLQPRTFLL | A*02:01 | None | 1 | 9.09314E-06 | 34341799 |
| 12 | CASSLGGNQPQHF | TRBV12-3 | TCRBV07-03*01 | LTDEMIAQY | A*01:01 | A*01:01:01 | 2 | 1.81863E-05 | 34341799 |
| 12 | CASSLGGNQPQHF | | TCRBV27-01*01 | | | | 1 | 9.09314E-06 | |
| 12 | CASSLGGNQPQHF | | TCRBV12-03/12-04*01 | | | | 18 | 0.000163677 | |
| 12 | CASSLGPEAFF | TRBV7-9 | TCRBV07-09*01 | YLQPRTFLL | A*02:01 | None | 1 | 9.09314E-06 | 34341799 |
| 12 | CASSLGPEAFF | | TCRBV05-04*01 | | | | 1 | 9.09314E-06 | |
| 12 | CASSLQNTGELFF | TRBV7-8 | TCRBV04-02*01 | YLQPRTFLL | A*02:01 | None | 1 | 9.09314E-06 | 33951417 |
| 12 | CASSLTSGAYNEQFF | TRBV19 | TCRBV05-01*01 | NQKLIANQF | B*15:01 | None | 1 | 9.09314E-06 | 34341799 |
| 12 | CASSLVVTGELFF | TRBV7-3 | TCRBV07-09*01 | LTDEMIAQY | A*01:01 | A*01:01:01 | 1 | 9.09314E-06 | 34341799 |
| 12 | CASSPDRNTGELFF | TRBV5-4 | TCRBV11-02*01 | YLQPRTFLL | A*02:01 | None | 1 | 9.09314E-06 | 33951417 |

| | | | | | | | | | |
|---|---|---|---|---|---|---|---|---|---|
| 12 | CASSPGQGYEQYF | TRBV6-4 | TCRBV27-01*01 | NYNYLYRLF | A*24:02 | None | 1 | 9.09314E-06 | 34341799 |
| 12 | CASSPGQGYEQYF | | TCRBV28-01*01 | | | | 1 | 9.09314E-06 | |
| 12 | CASSPGTSGGSSYNEQFF | TRBV11-2 | TCRBV05-01*01 | NQKLIANQF | B*15:01 | None | 2 | 1.81863E-05 | 34341799 |
| 12 | CASSQGGNEQYF | TRBV4-1 | TCRBV16-01*01 | YLQPRTFLL | A*02:01 | None | 1 | 9.09314E-06 | 34341799 |
| 12 | CASSQLGGYEQYF | TRBV4-3 | TCRBV04-01*01 | NYNYLYRLF | A*24:02 | None | 1 | 9.09314E-06 | 34341799 |
| 12 | CASSSQNTGELFF | TRBV7-8 | TCRBV05-01*01 | YLQPRTFLL | A*02:01 | None | 1 | 9.09314E-06 | 33326767 |
| 12 | CASSSQNTGELFF | | TCRBV11-02*03 | | | | 1 | 9.09314E-06 | |
| 12 | CASSVQNTGELFF | TRBV7-8 | TCRBV07-03*01 | YLQPRTFLL | A*02:01 | None | 1 | 9.09314E-06 | 33326767 |
| 12 | CASSYGNQPQHF | TRBV7-9 | TCRBV06-06*01 | YLQPRTFLL | A*02:01 | None | 1 | 9.09314E-06 | 34341799 |
| 12 | CSARDRSSYEQYF | TRBV20-1 | TCRBV20 | YLQPRTFLL | A*02:01 | None | 1 | 9.09314E-06 | 34341799 |
| 13 | CASSDLNTGELFF | TRBV2 | TCRBV02-01 | YLQPRTFLL | A*02:01 | A*02:01:01 | 1 | 1.42448E-05 | 33326767 |
| | | TRBV2 | | YLQPRTFLL | A*02:01 | A*02:01:01 | | | 33951417 |
| 13 | CASSEAGGYEQYF | TRBV2 | TCRBV07 | NYNYLYRLF | A*24:02 | A*24:02:01 | 1 | 1.42448E-05 | 34341799 |
| 13 | | TRBV6-1 | | NYNYLYRLF | A*24:02 | A*24:02:01 | | | 34341799 |
| 13 | CASSEGQGYEQYF | TRBV5-1 | TCRBV06-01*01 | NYNYLYRLF | A*24:02 | A*24:02:01 | 1 | 1.42448E-05 | 34341799 |
| | | TRBV2 | | NYNYLYRLF | A*24:02 | A*24:02:01 | | | 34341799 |
| | | TRBV6-1 | | NYNYLYRLF | A*24:02 | A*24:02:01 | | | 34341799 |
| 13 | CASSEMNTGELFF | TRBV2 | TCRBV28-01*01 | YLQPRTFLL | A*02:01 | A*02:01:01 | 1 | 1.42448E-05 | 33326767 |
| 13 | CASSFAVNTEAFF | TRBV27 | TCRBV11 | LTDEMIAQY | A*01:01 | None | 1 | 1.42448E-05 | 34341799 |
| 13 | CASSGTSGSTDTQYF | TRBV6-4 | TCRBV06-04*01 | NQKLIANQF | B*15:01 | None | 6 | 8.54689E-05 | 34341799 |
| 13 | CASSLAGYEQYF | TRBV5-1 | TCRBV07-08*01 | LTDEMIAQY | A*01:01 | None | 1 | 1.42448E-05 | 34341799 |
| 13 | CASSLAGYEQYF | | TCRBV07-09*01 | | | | 1 | 1.42448E-05 | |
| 13 | CASSLAVNTEAFF | TRBV27 | TCRBV07-08*01 | LTDEMIAQY | A*01:01 | None | 1 | 1.42448E-05 | 34341799 |
| 13 | CASSLAVNTEAFF | | TCRBV07-09*01 | | | | 1 | 1.42448E-05 | |
| 13 | CASSLGGNQPQHF | TRBV12 | TCRBV28-01*01 | LTDEMIAQY | A*01:01 | None | 1 | 1.42448E-05 | 34341799 |

| | | | | | | | | | |
|---|---|---|---|---|---|---|---|---|---|
| 13 | CASSLGGNQPQHF | | TCRBV13-01*01 | | | | 1 | 1.42448E-05 | |
| 13 | CASSLGPEAFF | TRBV7-9 | TCRBV07-09*01 | YLQPRTFLL | A*02:01 | A*02:01:01 | 1 | 1.42448E-05 | 34341799 |
| 13 | CASSLIGGSSGNTIYF | TRBV2 | TCRBV05-01*01 | QYIKWPWYI | A*24:02 | A*24:02:01 | 1 | 1.42448E-05 | 34341799 |
| 13 | CASSLVVTGELFF | TRBV7-3 | TCRBV07-09*01 | LTDEMIAQY | A*01:01 | None | 1 | 1.42448E-05 | 34341799 |
| 13 | CASSPGTSGGSSYNEQFF | TRBV11-2 | TCRBV18-01*01 | NQKLIANQF | B*15:01 | None | 1 | 1.42448E-05 | 34341799 |
| 13 | CASSPLSYEQYF | TRBV12-4 | TCRBV05-08*01 | LTDEMIAQY | A*01:01 | None | 2 | 2.84896E-05 | 34341799 |
| 13 | CASSSLNTGELFF | TRBV2 | TCRBV07-08*01 | YLQPRTFLL | A*02:01 | A*02:01:01 | 1 | 1.42448E-05 | 33326767 |
| 13 | CASSVAGSSYEQYF | TRBV2 | TCRBV02-01 | LTDEMIAQY | A*01:01 | None | 1 | 1.42448E-05 | 34341799 |
| 13 | CASSVAGSSYEQYF | | TCRBV06-02*02 | | | | 1 | 1.42448E-05 | |
| 13 | CASTGLNTGELFF | TRBV2 | TCRBV06-01*01 | YLQPRTFLL | A*02:01 | A*02:01:01 | 1 | 1.42448E-05 | 33326767 |
| 14 | CAISGGNEQFF | TRBV10-3 | TCRBV10-03*01 | QYIKWPWYI | A*24:02 | None | 1 | 8.97602E-06 | 34341799 |
| 14 | CASGQLNTGELFF | TRBV2 | TCRBV02-01 | YLQPRTFLL | A*02:01 | A*02:01:01 | 1 | 8.97602E-06 | 33951417 |
| | | TRBV7-8 | | YLQPRTFLL | A*02:01 | A*02:01:01 | | | 33951417 |
| 14 | CASSDLDTGELFF | TRBV2 | TCRBV02-01 | YLQPRTFLL | A*02:01 | A*02:01:01 | 1 | 8.97602E-06 | 33326767 |
| | | TRBV2 | | YLQPRTFLL | A*02:01 | A*02:01:01 | | | 34341799 |
| | | TRBV2 | | YLQPRTFLL | A*02:01 | A*02:01:01 | | | 33951417 |
| 14 | CASSDMNTGELFF | TRBV2 | TCRBV02-01 | YLQPRTFLL | A*02:01 | A*02:01:01 | 1 | 8.97602E-06 | 33326767 |
| 14 | CASSDTNTGELFF | TRBV5-1 | TCRBV06-04*01 | YLQPRTFLL | A*02:01 | A*02:01:01 | 2 | 1.7952E-05 | 33326767 |
| 14 | CASSELNTGELFF | TRBV7-8 | TCRBV25-01*01 | YLQPRTFLL | A*02:01 | A*02:01:01 | 1 | 8.97602E-06 | 33326767 |
| 14 | CASSESNTGELFF | TRBV10-1 | TCRBV10-01*01 | YLQPRTFLL | A*02:01 | A*02:01:01 | 1 | 8.97602E-06 | 33326767 |
| 14 | CASSFAVNTEAFF | TRBV27 | TCRBV07 | LTDEMIAQY | A*01:01 | None | 1 | 8.97602E-06 | 34341799 |
| 14 | CASSGGYEQYF | TRBV7-8 | TCRBV10-01*01 | YLQPRTFLL | A*02:01 | A*02:01:01 | 2 | 1.7952E-05 | 34341799 |
| 14 | CASSGGYEQYF | | TCRBV18-01*01 | | | | 1 | 8.97602E-06 | |

| | | | | | | | | | |
|---|---|---|---|---|---|---|---|---|---|
| 14 | CASSGGYEQYF | | TCRBV06 | | | | 1 | 8.97602E-06 | |
| 14 | CASSGGYEQYF | | TCRBV14-01*01 | | | | 1 | 8.97602E-06 | |
| 14 | CASSGTSGSTDTQYF | TRBV6-4 | TCRBV06-04*01 | NQKLIANQF | B*15:01 | None | 34 | 0.000305185 | 34341799 |
| 14 | CASSGTSGSTDTQYF | | TCRBV11-01*01 | | | | 1 | 8.97602E-06 | |
| 14 | CASSGTSGSTDTQYF | | TCRBV19-01*01 | | | | 1 | 8.97602E-06 | |
| 14 | CASSLAGYEQYF | TRBV5-1 | TCRBV07-06*01 | LTDEMIAQY | A*01:01 | None | 1 | 8.97602E-06 | 34341799 |
| 14 | CASSLAGYEQYF | | TCRBV28-01*01 | | | | 1 | 8.97602E-06 | |
| 14 | CASSLAGYEQYF | | TCRBV19-01*01 | | | | 1 | 8.97602E-06 | |
| 14 | CASSLAGYEQYF | | TCRBV12-03/12-04*01 | | | | 1 | 8.97602E-06 | |
| 14 | CASSLAGYEQYF | | TCRBV05-01*01 | | | | 1 | 8.97602E-06 | |
| 14 | CASSLAQGYEQYF | TRBV12-3 | TCRBV11-02*01 | NYNYLYRLF | A*24:02 | A*24:02:01 | 2 | 1.7952E-05 | 34341799 |
| 14 | CASSLASNQPQHF | TRBV27 | TCRBV05-05*01 | LTDEMIAQY | A*01:01 | None | 4 | 3.59041E-05 | 34341799 |
| 14 | CASSLAVNTEAFF | TRBV27 | TCRBV05-01*01 | LTDEMIAQY | A*01:01 | None | 1 | 8.97602E-06 | 34341799 |
| 14 | CASSLGASSYNEQFF | TRBV12-3 | TCRBV05-06*01 | YLQPRTFLL | A*02:01 | A*02:01:01 | 1 | 8.97602E-06 | 34341799 |
| 14 | CASSLGASSYNEQFF | | TCRBV11-02*01 | | | | 1 | 8.97602E-06 | |
| 14 | CASSLGGNQPQHF | TRBV12-3 | TCRBV28-01*01 | LTDEMIAQY | A*01:01 | None | 1 | 8.97602E-06 | 34341799 |
| 14 | CASSLGGNQPQHF | | TCRBV07-09*01 | | | | 1 | 8.97602E-06 | |
| 14 | CASSLGGNQPQHF | | TCRBV12-03/12-04*01 | | | | 1 | 8.97602E-06 | |
| 14 | CASSLQNTGELFF | TRBV7-8 | TCRBV05-01*01 | YLQPRTFLL | A*02:01 | A*02:01:01 | 1 | 8.97602E-06 | 33951417 |
| 14 | CASSLTSGAYNEQFF | TRBV19 | TCRBV07-06*01 | NQKLIANQF | B*15:01 | None | 1 | 8.97602E-06 | 34341799 |
| 14 | CASSNNYEQYF | TRBV19 | TCRBV07-07*01 | NYNYLYRLF | A*24:02 | A*24:02:01 | 1 | 8.97602E-06 | 34341799 |
| 14 | CASSPGQGYEQYF | TRBV6-4 | TCRBV12-03/12-04*01 | NYNYLYRLF | A*24:02 | A*24:02:01 | 1 | 8.97602E-06 | 34341799 |

| | | | | | | | | | |
|---|---|---|---|---|---|---|---|---|---|
| 14 | CASSPLSYEQYF | TRBV12-4 | TCRBV06-05*01 | LTDEMIAQY | A*01:01 | None | 1 | 8.97602E-06 | 34341799 |
| 14 | CASSPRTATYEQYF | TRBV7-8 | TCRBV19-01*01 | YLQPRTFLL | A*02:01 | A*02:01:01 | 1 | 8.97602E-06 | 34341799 |
| 14 | CASSQGQGNIQYF | TRBV4-3 | TCRBV03-01/03-02*01 | RLQSLQTYV | A*02:01 | A*02:01:01 | 1 | 8.97602E-06 | 33326767 |
| 14 | CASSSLNTGELFF | TRBV2 | TCRBV05-01*01 | YLQPRTFLL | A*02:01 | A*02:01:01 | 1 | 8.97602E-06 | 33326767 |
| 14 | CASSSSGTATYEQYF | TRBV7-9 | TCRBV28-01*01 | NQKLIANQF | B*15:01 | None | 1 | 8.97602E-06 | 34341799 |
| 14 | CASSTRDSNQPQHF | TRBV19 | TCRBV06-05*01 | YLQPRTFLL | A*02:01 | A*02:01:01 | 1 | 8.97602E-06 | 34341799 |
| 14 | CASSVDNTGELFF | TRBV9 | TCRBV06-04*01 | YLQPRTFLL | A*02:01 | A*02:01:01 | 1 | 8.97602E-06 | 33326767 |
| 14 | CASTGADTQYF | TRBV6-6 | TCRBV02-01 | YLQPRTFLL | A*02:01 | A*02:01:01 | 1 | 8.97602E-06 | 34341799 |
| 15 | CASSDSYGYTF | TRBV7-8 | TCRBV06-04*01 | YLQPRTFLL | A*02:01 | None | 1 | 1.55272E-05 | 33326767 |
| | | TRBV7-8 | | YLQPRTFLL | A*02:01 | | | | 33951417 |
| 15 | CASSEGQGYEQYF | TRBV5-1 | TCRBV02-01 | NYNYLYRLF | A*24:02 | None | 1 | 1.55272E-05 | 34341799 |
| | | TRBV2 | | NYNYLYRLF | A*24:02 | | | | 34341799 |
| | | TRBV6-1 | | NYNYLYRLF | A*24:02 | | | | 34341799 |
| 15 | CASSGGYEQYF | TRBV7-8 | TCRBV27-01*01 | YLQPRTFLL | A*02:01 | None | 1 | 1.55272E-05 | 34341799 |
| | CASSGGYEQYF | | TCRBV06-06 | | | | 1 | 1.55272E-05 | |
| 15 | CASSGTSGSTDTQYF | TRBV6-4 | TCRBV06-04*01 | NQKLIANQF | B*15:01 | None | 2 | 3.10545E-05 | 34341799 |
| 15 | CASSLAGPNEQFF | TRBV7-9 | TCRBV05-06*01 | YLQPRTFLL | A*02:01 | None | 1 | 1.55272E-05 | 33951417 |
| 15 | CASSLGASSYNEQFF | TRBV12-3 | TCRBV07-03*01 | YLQPRTFLL | A*02:01 | None | 1 | 1.55272E-05 | 34341799 |
| 15 | CASSLGGNQPQHF | TRBV12-3 | TCRBV07-02 | LTDEMIAQY | A*01:01 | A*01:01:01 | 2 | 3.10545E-05 | 34341799 |
| 15 | CASSLGGNQPQHF | | TCRBV07-06*01 | | | | 1 | 1.55272E-05 | |
| 15 | CASSLQNTGELFF | TRBV7-8 | TCRBV05-05*01 | YLQPRTFLL | A*02:01 | None | 1 | 1.55272E-05 | 33951417 |
| 15 | CASSLTGGGYTF | TRBV12-4 | TCRBV27-01*01 | NQKLIANQF | B*15:01 | None | 1 | 1.55272E-05 | 34341799 |
| 15 | CASSNSYGYTF | TRBV7-8 | TCRBV06-05*01 | YLQPRTFLL | A*02:01 | None | 1 | 1.55272E-05 | 33326767 |

| | | | | | | | | | |
|---|---|---|---|---|---|---|---|---|---|
| 15 | CASSPDRNTGELFF | TRBV5-4 | TCRBV06-04*01 | YLQPRTFLL | A*02:01 | None | 1 | 1.55272E-05 | 33951417 |
| 15 | CASSPGQGYEQYF | TRBV6-4 | TCRBV18-01*01 | NYNYLYRLF | A*24:02 | None | 1 | 1.55272E-05 | 34341799 |
| 15 | CASSTGNQPQHF | TRBV9 | TCRBV18-01*01 | AEVQIDRLI | B*44:02 | None | 1 | 1.55272E-05 | 34341799 |
| 15 | CASSTRDSNQPQHF | TRBV19 | TCRBV09-01*01 | YLQPRTFLL | A*02:01 | None | 1 | 1.55272E-05 | 34341799 |
| 15 | CASSVAGSSYEQYF | TRBV2 | TCRBV09-01*01 | LTDEMIAQY | A*01:01 | A*01:01:01 | 1 | 1.55272E-05 | 34341799 |
| 16 | CAISESSGYEQYF | TRBV10-3 | TCRBV10-03*01 | NYNYLYRLF | A*24:02 | A*24:02:01 | 1 | 1.05945E-05 | 34341799 |
| 16 | CASSEANTGELFF | TRBV7-9 | TCRBV06-01*01 | YLQPRTFLL | A*02:01 | A*02:01:01 | 1 | 1.05945E-05 | 33326767 |
| 16 | CASSEANTGELFF | TRBV7-9 | TCRBV02-01 | YLQPRTFLL | A*02:01 | A*02:01:01 | 1 | 1.05945E-05 | 33326767 |
| 16 | CASSESNTGELFF | TRBV10-1 | TCRBV25-01*01 | YLQPRTFLL | A*02:01 | A*02:01:01 | 1 | 1.05945E-05 | 33326767 |
| 16 | CASSETGGYEQYF | TRBV6-4 | TCRBV10-01*01 | NYNYLYRLF | A*24:02 | A*24:02:01 | 2 | 2.11889E-05 | 34341799 |
| | | TRBV6-1 | | NYNYLYRLF | A*24:02 | A*24:02:01 | | | 34341799 |
| | | TRBV4-1 | | NYNYLYRLF | A*24:02 | A*24:02:01 | | | 34341799 |
| 16 | CASSGGYEQYF | TRBV7-8 | TCRBV02-01 | YLQPRTFLL | A*02:01 | A*02:01:01 | 1 | 1.05945E-05 | 34341799 |
| 16 | CASSGGYEQYF | | TCRBV06-05*01 | | | | 1 | 1.05945E-05 | |
| 16 | CASSGTSGSTDTQYF | TRBV6-4 | TCRBV06-04*01 | NQKLIANQF | B*15:01 | B*15:01:01 | 5 | 5.29723E-05 | 34341799 |
| 16 | CASSLAGYEQYF | TRBV5-1 | TCRBV07-06*01 | LTDEMIAQY | A*01:01 | None | 1 | 1.05945E-05 | 34341799 |
| 16 | CASSLAGYEQYF | | TCRBV05-08*01 | | | | 1 | 1.05945E-05 | |
| 16 | CASSLAQGYEQYF | TRBV12-3 | TCRBV05-06*01 | NYNYLYRLF | A*24:02 | A*24:02:01 | 1 | 1.05945E-05 | 34341799 |
| 16 | CASSLASNQPQHF | TRBV27 | TCRBV27-01*01 | LTDEMIAQY | A*01:01 | None | 2 | 2.11889E-05 | 34341799 |
| 16 | CASSLEDTNYGYTF | TRBV7-2 | TCRBV07-02 | NQKLIANQF | B*15:01 | B*15:01:01 | 2 | 2.11889E-05 | 34341799 |
| 16 | CASSLEGIGGYTF | TRBV7-6 | TCRBV28-01*01 | LTDEMIAQY | A*01:01 | None | 1 | 1.05945E-05 | 34341799 |
| 16 | CASSLGGNQPQHF | TRBV12-3 | TCRBV05-01*01 | LTDEMIAQY | A*01:01 | None | 1 | 1.05945E-05 | 34341799 |
| 16 | CASSLGGNQPQHF | | TCRBV07-08*02 | | | | 1 | 1.05945E-05 | |
| 16 | CASSLGGNQPQHF | | TCRBV27-01*01 | | | | 1 | 1.05945E-05 | |

| | | | | | | | | | |
|---|---|---|---|---|---|---|---|---|---|
| 16 | CASSLGGNQPQHF | | TCRBV07-09*01 | | | | 1 | 1.05945E-05 | |
| 16 | CASSLGIAKNIQYF | TRBV11-2 | TCRBV11-02*03 | YLQPRTFLL | A*02:01 | A*02:01:01 | 2 | 2.11889E-05 | 34341799 |
| 16 | CASSLGISGELFF | TRBV7-8 | TCRBV05-01*01 | YLQPRTFLL | A*02:01 | A*02:01:01 | 1 | 1.05945E-05 | 33951417 |
| 16 | CASSLGPEAFF | TRBV7-9 | TCRBV11-02*03 | YLQPRTFLL | A*02:01 | A*02:01:01 | 1 | 1.05945E-05 | 34341799 |
| 16 | CASSNRGGNEQFF | TRBV19 | TCRBV19-01*01 | NQKLIANQF | B*15:01 | B*15:01:01 | 2 | 2.11889E-05 | 34341799 |
| 16 | CASSPEDTQYF | TRBV7-9 | TCRBV18-01*01 | YLQPRTFLL | A*02:01 | A*02:01:01 | 1 | 1.05945E-05 | 34341799 |
| 16 | | TRBV7-9 | | YLQPRTFLL | A*02:01 | A*02:01:01 | | | 33326767 |
| 16 | CASSPGTTLAKNIQYF | TRBV7 | TCRBV06 | YLQPRTFLL | A*02:01 | A*02:01:01 | 3 | 3.17834E-05 | 33664060 |
| 16 | CASSQGGNEQYF | TRBV4-1 | TCRBV03-01/03-02*01 | YLQPRTFLL | A*02:01 | A*02:01:01 | 1 | 1.05945E-05 | 34341799 |
| 16 | CASSQGPNGYTF | TRBV4-1 | TCRBV03-01/03-02*01 | YLQPRTFLL | A*02:01 | A*02:01:01 | 1 | 1.05945E-05 | 34341799 |
| 16 | CASSTGGRGEQFF | TRBV7-3 | TCRBV28-01*01 | LTDEMIAQY | A*01:01 | None | 1 | 1.05945E-05 | 34341799 |
| 16 | CASSVAGSSYEQYF | TRBV2 | TCRBV06 | LTDEMIAQY | A*01:01 | None | 1 | 1.05945E-05 | 34341799 |
| 16 | CASSYGNQPQHF | TRBV7-9 | TCRBV06-02/06-03*01 | YLQPRTFLL | A*02:01 | A*02:01:01 | 1 | 1.05945E-05 | 34341799 |
| 16 | CASTSLNTGELFF | TRBV2 | TCRBV10-02*01 | YLQPRTFLL | A*02:01 | A*02:01:01 | 1 | 1.05945E-05 | 33326767 |
| 16 | CAWSVGGNYGYTF | | TCRBV30-01*01 | SKRSFIEDL LFNKVTLA | DPA1*01:03/ DPB1*04:01 | Unknown | 1 | 1.05945E-05 | 33830946 |

**Table S4. PBMC-derived TCR sequences that match previously published sequences obtained from SARS2 Spike-loaded tetramer or multimer experiments are shown.** Green highlight indicates identical V gene usage between the published sequence and the PBMC-derived sequence found in this study. Blue highlight indicates concordance between previously published HLA epitope restriction and the participant's HLA type.

| Participant | Breastmilk-derived CDR3 amino acid sequence | Clustered reference CDR3 amino acid sequence | TCRdist units | Cognate Spike-specific epitope of reference receptor | HLA restriction of epitope (reference) | Relevant HLA allele of participant (this study) | Templates (this study) | Productive frequency (this study) | PMID of reference sequence |
|---|---|---|---|---|---|---|---|---|---|
| 3 | CASSLDIEQFF | CASSLDIEQYF | -1 | YLQPRTFLL | A*02:01 | Typing not available, presumed A*02:01 | 1 | 0.000318 | 34341799<br>33951417 |
| 16 | CASSITSGAYNEQFF | CASSLTSGAYNEQFF | 6 | NQKLIANQF | B*15:01 | B*15:01:01 | 3 | 0.000613 | 34341799 |
| 3 | CASSLDIQAFF | CASSLDIEAFF | 6 | YLQPRTFLL | A*02:01 | Typing not available, presumed A*02:01 | 17 | 0.00541 | 33326767<br>34341799<br>33951417 |
| 16 | CSVEVADYEQYF | CSVEVSDYEQYF | 9 | NQKLIANQF | B*15:01 | B*15:01:01 | 1 | 0.00020 | 34341799 |
| 12 | CASSEASGYEQYF | CASSEATGYEQYF | 9 | NYNYLYRLF | A*24:02 | None | 2 | 0.00196 | 34341799 |
| 3 | CASSLDIQAFF | CASSLDIKAFF | 9 | YLQPRTFLL | A*02:01 | Typing not available, presumed A*02:01 | 17 | 0.00541 | 34341799 |

**Table S5. Previously published sequences obtained from SARS2 Spike-loaded tetramer or multimer experiments were used in conjunction with the tcrdist3 algorithm to predict novel Spike-specific T cell clones in breastmilk of study participants.**

| Marker | Clone | Fluorochrome | Manufacturer | Catalog # | Dilution |
|---|---|---|---|---|---|
| CD45 | 2D1 | FITC | BioLegend | 368508 | 1:80 |
| CD3 | SK7 | PE-Cy7 | BD | 557851 | 1:80 |
| CD4 | RPA-T4 | APC-Cy7 | BD | 560158 | 1:80 |
| CD8 | RPA-T8 | BB700 | BD | 566452 | 1:100 |
| CCR9 | L053E8 | BV421 | BioLegend | 358914 | 1:50 |
| CCR7 | 3D12 | PE | BD | 552176 | 1:50 |
| CD45RO | UCHL1 | APC | BioLegend | 304210 | 1:80 |
| CD103 | Ber-ACT8 | BV786 | BioLegend | 350230 | 1:50 |
| LIVE/DEAD™ Fixable Aqua | | | Invitrogen | L34957 | 1:1000 |

**Table S6. Flow cytometry panel 1.**

| Marker | Clone | Fluorochrome | Manufacturer | Catalog # | Dilution |
|---|---|---|---|---|---|
| Spike-loaded tetramer | | APC | | | 1:100 |
| CD25 | 2A3 | BUV737 | BD | 612807 | 1:50 |
| α4β7 | FAB10078U | AF350 | R&D | FAB10078U-100UG | 1:40 |
| CD103 | Ber-ACT8 | BV786 | BioLegend | 350230 | 1:50 |
| CCR7 | G043H7 | BV711 | BioLegend | 353228 | 1:50 |
| CD127 | A019D5 | BV650 | BioLegend | 351326 | 1:80 |
| CD69 | FN50 | BV605 | BioLegend | 310938 | 1:50 |
| CCR9 | L053E8 | BV421 | BioLegend | 358914 | 1:50 |
| CD8 | RPA-T8 | BB700 | BD | 566452 | 1:100 |
| CCR5 | 3A9 | FITC | BD | 564512 | 1:40 |
| CD3 | SK7 | PE-Cy7 | BD | 557851 | 1:80 |
| CXCR4 | 12G5 | PE-Cy5 | BioLegend | 309508 | 1:40 |
| CD45RO | UCHL1 | PE-CF594 | BD | 562299 | 1:80 |
| CD137 | 4B4-1 | PE | BD | 555956 | 1:50 |
| CD4 | RPA-T4 | APC-H7 | BD | 560158 | 1:80 |
| CD45 | 2D1 | AF700 | BioLegend | 368514 | 1:80 |
| CD14 | MHCD1418 | PE-Cy5.5 | Life Technologies | MHCD1418 | 1:160 |
| LIVE/DEAD™ Fixable Aqua | | | Invitrogen | L34957 | 1:1000 |

**Table S7. Flow cytometry panel 2.** Cells were stained with Spike epitope-loaded tetramer, washed, and then stained with the remainder of the panel.

| Antibody of Interest | Dilution of Secondary Antibody | Development Time (minutes) | Initial Plasma Dilution per Antibody |
|---|---|---|---|
| Total IgG* | 1:4000 | 2.5 – 3.5 | 1:20 |
| IgG1 | 1:1000 | 10 | 1:2 |
| IgG3 | 1:1000 | 10 | 1:2 |
| IgG4 | 1:2000 | 10 | 1:2 |
| IgM | 1:2000 | 3.5 – 4.5 | 1:20 |
| IgA | 1:10000 | 3.5 – 4.5 | 1:20 |
| *RBD qualification was only completed for total IgG. | | | |

**Table S8. Parameters for each immunoglobulin qualified in ELISA methodology.**
